# Circulating Proteome for Pulmonary Nodule Malignancy

**DOI:** 10.1101/2022.09.24.22280288

**Authors:** Elham Khodayari Moez, Yonathan Brhane, Matthew Warkentin, Stephen Lam, John K Field, Geoffrey Liu, Luis M Montuenga, Javier J Zulueta, Karmele Valencia, Miguel Mesa-Guzman, Sukhinder Atkar-Khattra, Michael PA Davies, Benjamin Grant, Andrea Pasquier Nialet, Christopher I Amos, Hilary A Robbins, Mattias Johansson, Rayjean J Hung

**Author notes:** **Corresponding author** Rayjean Hung, Prosserman Centre for Population Health Research, Lunenfeld-Tanenbaum Research Institute, Sinai Health, Dalla Lana School of Public Health, University of Toronto, 60 Murray St. Toronto, ON M5T 3L9. Canada. **Disclaimer** Where authors are identified as personnel of the International Agency for Research on Cancer / World Health Organization, the authors alone are responsible for the views expressed in this article and they do not necessarily represent the decisions, policy or views of the International Agency for Research on Cancer / World Health Organization. **Ethics** Ethics committee/IRB of the Mount Sinai Hospital gave ethical approval for this work. The Ethics approval was also obtained from local institutes at each study sites. **Availability of Data** The dataset presented in this study is available from the corresponding author upon request and committee approval.

## Abstract

**Background:** While lung cancer low-dose computed tomography (LDCT) screening is being rolled out in many regions around the world, differentiation of indeterminate pulmonary nodules between malignant and benign remains to a challenge for screening programs. We conducted one of the first systematic investigations of circulating protein markers for their ability to assess the risk of malignancy for screen-detected pulmonary nodules.

**Methods:** Based on four LDCT screening studies in the United States, Canada and Europe, we assayed 1078 unique protein markers in pre-diagnostic samples based on a nested case-control design with a total of 1253 participants. Protein markers were measured using proximity extension assays and the data were analyzed using multivariate logistic regression, random forest, and penalized regressions.

**Results:** We identified 36 potentially informative markers differentiating malignant nodules from benign nodules. Pathway analysis revealed a tightly connected network based on the 36 protein-coding genes. We observed a differential mRNA expression profile of the corresponding 36 mRNAs between lung tumors and adjacent normal tissues using data from The Cancer Genomic Atlas. We prioritized a panel of 9 protein markers through 10-fold nested cross-validations. We observed that circulating protein markers can increase sensitivity to 0.80 for nodule malignancy compared to the Brock model (p-value<0.001). Two additional markers were identified that were specific for lung tumors diagnosed within one year. All 11 protein markers showed general consistency in improving prediction across the four LDCT studies.

**Conclusions:** Circulating protein markers can help to differentiate between malignant and benign pulmonary nodules. Validating these results in an independent CT-screening study will be required prior to clinical implementation.

## INTRODUCTION

Lung cancer continues to represent a significant public health burden worldwide as the leading cause of global cancer death [1-3]. Low-dose computed tomography (LDCT) screening in smokers was shown to reduce lung cancer-related mortality by 20-30% [4-6], and as a result, the United States Preventive Services Task Force (USPSTF) recommends annual LDCT screening for those aged 50 to 80 years with at least 20 packyears smoking history, who either currently smoke or quit within the last 15 years [7]. However, challenges of implementing LDCT at the population-level remain. One of the main challenges is that pulmonary abnormalities can be found in approximately 20% of the screening participants, but only a small fraction of these nodules are actually malignant [6, 8].

The Lung-RADS (Lung CT Screening Reporting and Data System) classification system developed by the American College of Radiology is commonly used in the US screening program [9], but it is largely based on nodule diameters and solidity without considering other parameters, and currently there is still a wide range of nodule management protocols [8, 10, 11]. To differentiate benign from malignant pulmonary non-calcified nodules, several malignancy probability models have been proposed [12-15]. For example, the Brock model combines demographic, clinical and specific nodule information to assess the probability of nodule malignancy [15] and was incorporated into the British Thoracic Society guidelines [16]. While beneficial for reducing false positives, these nodule malignancy assessment tools are highly dependent on nodule sizes and have suboptimal predictive performance in small nodules.

Given the clinical importance of accurately classifying nodules based on their probability of malignancy in order to minimize any unnecessary follow-up procedures, we launched a systematic investigation of whether the circulating proteins can help to differentiate benign from malignant nodules in conjunction with the nodule features, and an individual’s medical history. Circulating protein markers can be ideal biomarkers, as they have been shown to predict cancer occurrence in prospective studies and can be obtained via a minimally invasive approach [17, 18]. While there have been a plethora of biomarker studies on lung cancer risk, the data on pulmonary nodule malignancy are much more sparse, and often limited to either a few targeted markers or a single study [19]. This is the first study to apply an extensive circulating proteomic approach using pre-clinical diagnostic samples based on international collaborations of four LDCT studies. Our goal was to identify circulating protein markers that can help to differentiate malignant from benign pulmonary nodules, beyond what can be achieved by the predictors included in the established Brock model.

## METHODS

### Study Design and Participants

As part of the Integrative Analysis of Lung Cancer Risk and Etiology (INTEGRAL) research program, this study was conducted based on four LDCT lung cancer screening programs in Canada, Spain, USA and UK using a nested case-control design, including the Pan-Canadian Early Detection of Lung Cancer Study (PanCan), the UK Lung Screening Trial (UKLS), the Toronto International Early Lung Cancer Action Program (IELCAP-Toronto) and the Pamplona International Early Lung Cancer Action Program (P-IELCAP) [20]. Detailed study designs for these studies have been previously reported [21-26]. In brief, varying by individual studies, these CT screening studies were conducted between 2001 to 2020 based on heavy and recent smokers aged 40 to 84, and the participants were followed for up to 17 years after enrollment. Participants completed a study questionnaire at baseline, including demographics, family history of lung cancer, personal history of cancer and history of COPD/emphysema. Blood samples were collected at baseline and at some of the follow-up screening rounds, depending on the study design (**Supplementary Methods**).

Patients with confirmed lung cancer diagnosis within 5 years after pre-diagnostic blood sample collection were included in this study as the case group. The study participants with benign pulmonary nodules but did not develop lung cancer at any time point including the follow-up period constituted the benign nodule control group. Cases and controls were frequency-matched on sex, age at enrollment, age at the abnormal finding, age at blood collection, and follow-up time. When multiple nodule controls were available, we selected the nodule with the highest probability of malignancy based on Brock model [12]. A total of 425 lung cancer cases and 430 frequency-matched controls with benign nodules were included in this analysis. In addition to controls with benign nodules, we selected 398 healthy controls without any nodules based on frequency matching with cases on age of enrollment, sex and follow up time. The healthy controls were not included in this specific analysis *per se* but were used to provide information for background protein expression distribution in the study source population.

The nodule features were recorded by the study radiologists, including the number of detected nodules, the nodule sizes, locations, type (solid, non-solid and part-solid) and the presence or absence of spiculations. The lungRADs category for each nodule was assigned by a radiologist. Written informed consent was obtained from all participants. The ethics approvals were obtained by each local institute and the Mount Sinai Hospital Research Ethics Board.

### Protein Biomarkers

The circulating proteome in pre-diagnostic plasma samples was quantified using Proximity Extension Assay (PEA), in which antibody-based immunoassay proteins was quantified by PCR extension when antibody-linked oligonucleotides are brough into close proximity [27]. A total of 1105 PEA assays representing 1078 unique protein markers were measured for a total of 1253 subjects using 12 panels, including panels of oncology, cardiovascular disease, inflammation, and metabolism assays (**Supplementary Table 1**). The relative abundance of protein levels was measured as Normalized Protein eXpression (NPX) values, which was calculated from the inverse amount of target nucleic acid based on the cycle threshold (Ct) values and expressed in the log_2_ scale. In each sample, internal controls were added to monitor three key assay steps, including incubation, extension and amplification/detection. In each plate, inter-plate controls, negative controls and sample controls were added to monitor variations between assays and background noise, and based on which, NPX values were normalized to minimize both intra and inter-assay variations [27]. The sample plates passed the quality control (QC) if the standard deviation of the internal control samples did not exceed 0.2 NPX. The individual samples passed the QC if the value for internal spiked controls deviated no more than 0.3 NPX from the plate median. The NPX values that did not pass QC were excluded from the analysis.

### Data Analyses

The NPX values for each CT screening cohort were standardized by z-transformation before performing pooled analysis across the four studies. To assess the association between each protein maker and nodule malignancy, while accounting for nodule characteristics, we applied multi0variate logistic regression adjusting for the Brock Score, which was computed based on age, sex, family history of lung cancer, emphysema, nodule size, nodule type, nodule location and nodule count as previously described [12]. For individuals with multiple nodules, the highest Brock Score was included in the model as it represents the nodule with the highest probability of developing into lung cancer. As different protein markers may have different timing of detectability before cancer diagnosis, we conducted stratified analysis by the time intervals between the time of blood test (corresponding to when the blood sample was collected) and cancer diagnosis, as less than 1 year, 1 to 3 years and more than 3 years, with a specific aim to identify protein markers that are most informative for imminent lung cancer (e.g. diagnosed within 1 year) and to assess if the protein markers remain informative with longer lead time.

To select the top markers that can potentially inform nodule malignancy, two analytical methods – penalized regression (LASSO) and random forest-were performed in parallel on the 1078 protein markers [28, 29]. Using LASSO with 500 random samplings of 80% of the data, the markers were considered potentially informative if selected by LASSO in at least 50% of the resamples; which resulted in selecting 25 markers. Using random forest, the markers were ranked by their importance values which reflects their relevance in classifying cases vs controls[30]. To allow both analytical methods to be equally considered, we selected the top 25 markers based on the importance values from random forest analysis. With 14 markers selected by both LASSO and random forest, a total of 36 markers were considered to be informative for nodule malignancy. The overview of our analytical pipeline is outlined in **Figure 1**.

**Figure 1:**
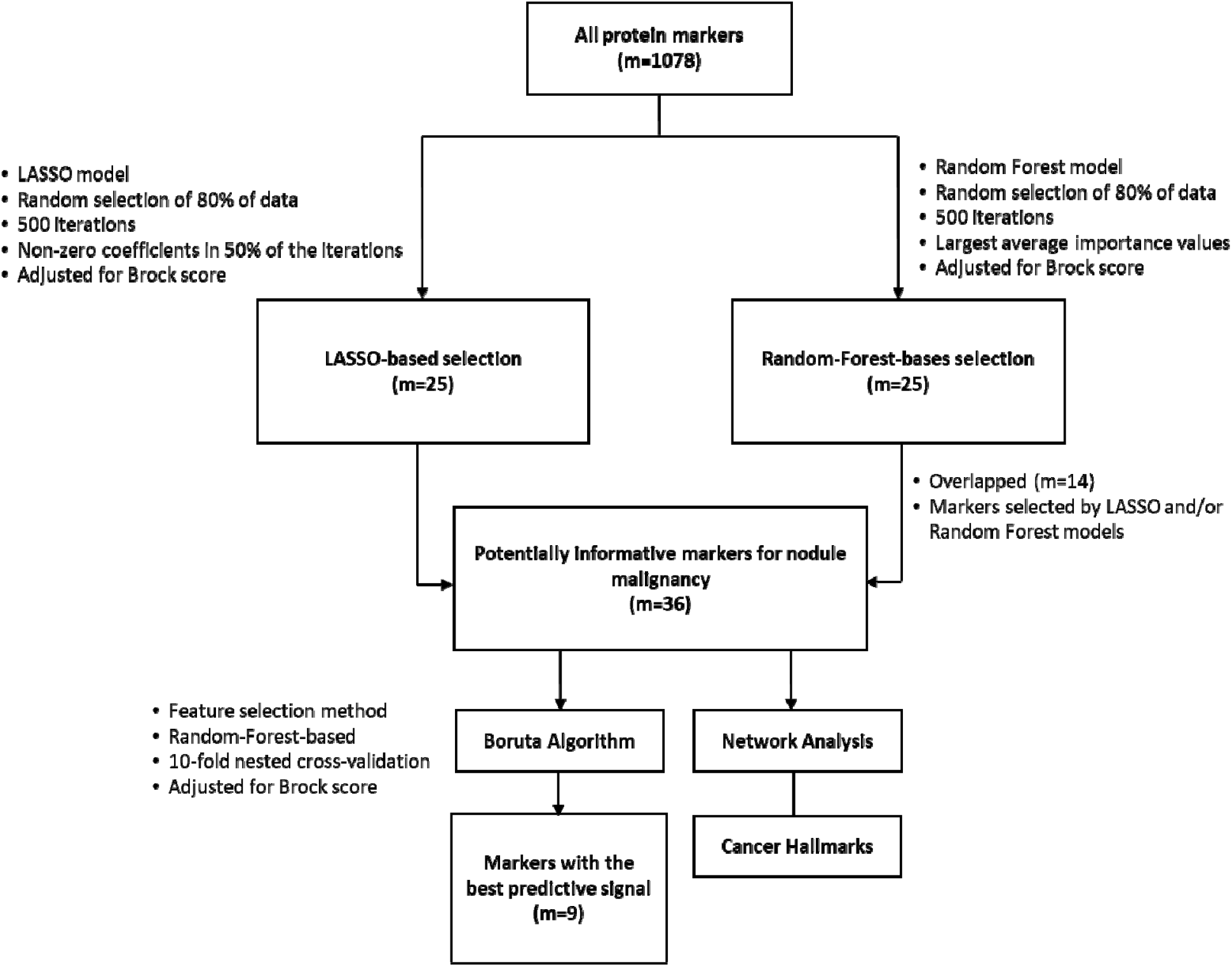
The overall analytical pipeline. LASSO, Least Absolute Shrinkage and Selection Operator. m = no. of markers

#### Pathway Enrichment and Network Analysis

To assess if any biological pathways were enriched in this selected set of 36 informative protein markers, we conducted the pathway enrichment analyses using Gene Ontology – Biological Process (GO-BP) pathway [31], focusing on the pathways in level 2 and level 3. We assessed if any pathway of interest contained more informative markers than one would expect just by chance based on the hypergeometric test. The pathways with False Discovery Rate (FDR)-adjusted p-values less than 0.01 were considered to be significantly enriched [32]. The fold enrichment values were calculated to quantify the degree of pathway over-representation [33].

#### Classification Accuracy

To narrow down the top protein markers for nodule differentiation, we conducted feature selection using the Boruta algorithm, a random forest-based approach [34]. We opted for a random forest approach for this step because random forests models account for non-linearities and interactions, and is considered a better suited model when the ultimate goal is to identify a combination of markers[35]. Brock score was included in the model to assess the added value of the circulating protein markers, and it was computed as the linear combination of age, sex, family history of lung cancer, emphysema and nodule features including the size, type, count, location and spiculation status of the nodules as previously described [15]. We applied a 10-fold nested cross-validation procedure: The inner layer was used for selecting the best model; the outer layer was used to evaluate the model performance using the specific segment of the data not used for training, and this process was repeated 10 times for each fold. The model that had the best overall classification accuracy when fitted in a random forest model was then further evaluated in the subset by nodule size, lead time to cancer diagnoses and histology groups. To find the maximum classification potential of the models, the sensitivity and specificity values were estimated based on the threshold corresponding to the largest Youden’s Index [36], or clinically meaningful threshold based on LungRADs categories, where LungRADs category 4 was assumed to predict the malignant nodules.

#### Differential protein expression profiles based on TCGA data

To evaluate if the expression profiles of these protein markers are indeed differential between lung tumor and lung normal tissues in an independent dataset, we applied unsupervised hierarchical clustering to the lung tissue RNAseq data in The Cancer Genome Atlas (TCGA) dataset [37, 38], including 537 lung adenocarcinoma with 59 adjacent lung normal tissues and 502 squamous cell carcinoma with 49 adjacent lung normal tissues.

## RESULTS

The key characteristics of CT screening participants included in this project by study are summarized in **Table 1**. The participants with malignant nodules (cases) and benign nodules (controls) groups had comparable distribution of age and sex from frequency-matching. The case group had higher proportion of the current smokers and was more likely to have a history of emphysema/COPD. As expected, the malignant nodules were on average larger than benign lesions. Adenocarcinoma is the predominant histological type among lung cancers diagnosed.

**Table 1:** Key characteristics of the study population by nodule malignancy status and CT screening cohorts.

|  |  | Lung cancer case |  |  |  |  | Benign nodule control |  |  |  |  | Total |
| --- | --- | --- | --- | --- | --- | --- | --- | --- | --- | --- | --- | --- |
|  |  | IELCAP-Toronto | PanCan | IELCAP-Pamplona | UKLS | Total | IELCAP-Toronto | PanCan | IELCAP-Pamplona | UKLS | Total |  |
| Total |  | 79 | 169 | 76 | 101 | 425 | 87 | 169 | 82 | 92 | 430 | 855 |
| Age at enrollment | mean (*SD*) | 62.7 (7) | 63.4 (6) | 63.8 (89) | 68.6 (4) | 64.6 (7) | 64.5 (7) | 64.3 (6) | 62.7 (9) | 68.5 (4) | 64.9 (7) | 64.8 (7) |
| Male | n (%) | 23 (29) | 83 (49) | 63 (83) | 80 (79) | 249 (59) | 25 (29) | 82 (48) | 66 (80) | 70 (76) | 243 (56) | 492 (57) |
| Current smokers | n (%) | 49 (62) | 109 (64) | 47 (62) | 60 (59) | 265 (62) | 30 (34) | 98 (58) | 44 (54) | 51 (55) | 223 (52) | 488 (57) |
| Histology |  |  |  |  |  |  |  |  |  |  |  |  |
| Adenocarcinoma | n (%) | 46 (58) | 97 (57) | 36 (47) | 48 (47) | 227 (53) | - | - | - | - | - | - |
| Squamous cell | n (%) | 7 (89) | 21 (12) | 15 (20) | 26 (26) | 69 (16) | - | - | - | - | - | - |
| Small cell | n (%) | 2 (2) | 11 (6) | 5 (7) | 10 (10) | 28 (7) | - | - | - | - | - | - |
| Stage |  |  |  |  |  |  |  |  |  |  |  |  |
| Stage 1 | n (%) | 59 (81) | 109 (70) | 57 (78) | 49 (50) | 274 (68) | - | - | - | - | - | - |
| Stage 2 | n (%) | 5 (7) | 9 (6) | 3 (4) | 9 (9) | 26 (6) | - | - | - | - | - | - |
| Stage 3 | n (%) | 4 (5) | 21 (13) | 7 (10) | 14 (14) | 46 (11) | - | - | - | - | - | - |
| Stage 4 | n (%) | 5 (7) | 17 (11) | 6 (8) | 26 (27) | 54 (13) | - | - | - | - | - | - |
| Nodule Size | median (*IQR*) | 12.0 (11) | 11.0 (10) | 10.0 (6) | 18.5 (13) | 12.0 (11) | 7.0 (2) | 4.9 (4) | 7.0 (12) | 6.9 (4) | 6.1 (4) | 8 (7) |
| Emphysema/COPD | n (%) | 16 (20) | 36 (21) | 62 (82) | 31 (31) | 145 (34) | 14 (16) | 31 (18) | 46 (56) | 20 (22) | 111 (26) | 256 (30) |
| Family history of lung cancer | n (%) | 13 (16) | 64 (38) | - | 23 (23) | 100 (23) | 15 (17) | 49 (29) | - | 22 (24) | 86 (20) | 186 (22) |

### The association of single protein markers

The single-marker results of all 1078 circulating protein markers are shown in the **Figure 2**, with odds ratios (OR) and significance level derived from the multivariate logistic regression. The ORs of the individual markers ranged from 0.75 to 1.46 after adjusting for the Brock score. The most significant protein markers are WAP four-disulfide core domain 2 (WFDC2) (OR= 1.49, 95%CI=1.26-1.76, p<0.001), tumor necrosis factor-related apoptosis-inducing ligand-receptor 2 (TRAIL-R2) (OR=1.46, 95%CI=1.22-1.76, p<0.001) and C-X-C motif chemokine ligand 17 (CXCL17) (OR= 1.42, 95%CI=1.19-1.70, p<0.001). Some of the markers showed an inverse association with nodule malignancy, for example fas ligand (FASLG) with OR of 0.77 (95%CI=0.66-0.90, p<0.001). In addition, keratin 19 (KRT19), matrix metalloproteinases 12 (MMP12) and Carcinoembryogenic antigen cell adhesion molecule 5 (CEACAM5) associated with lung cancer diagnosed within 1 year of lead time, with OR of 1.84 (95%CI=1.43-2.38, p<0.001), 1.60 (95%CI=1.27-2.04, p<0.001), and 1.43 (95%CI=1.13-1.83, p=0.002), respectively (**Supplementary Figure 1**).

**Figure 2:**
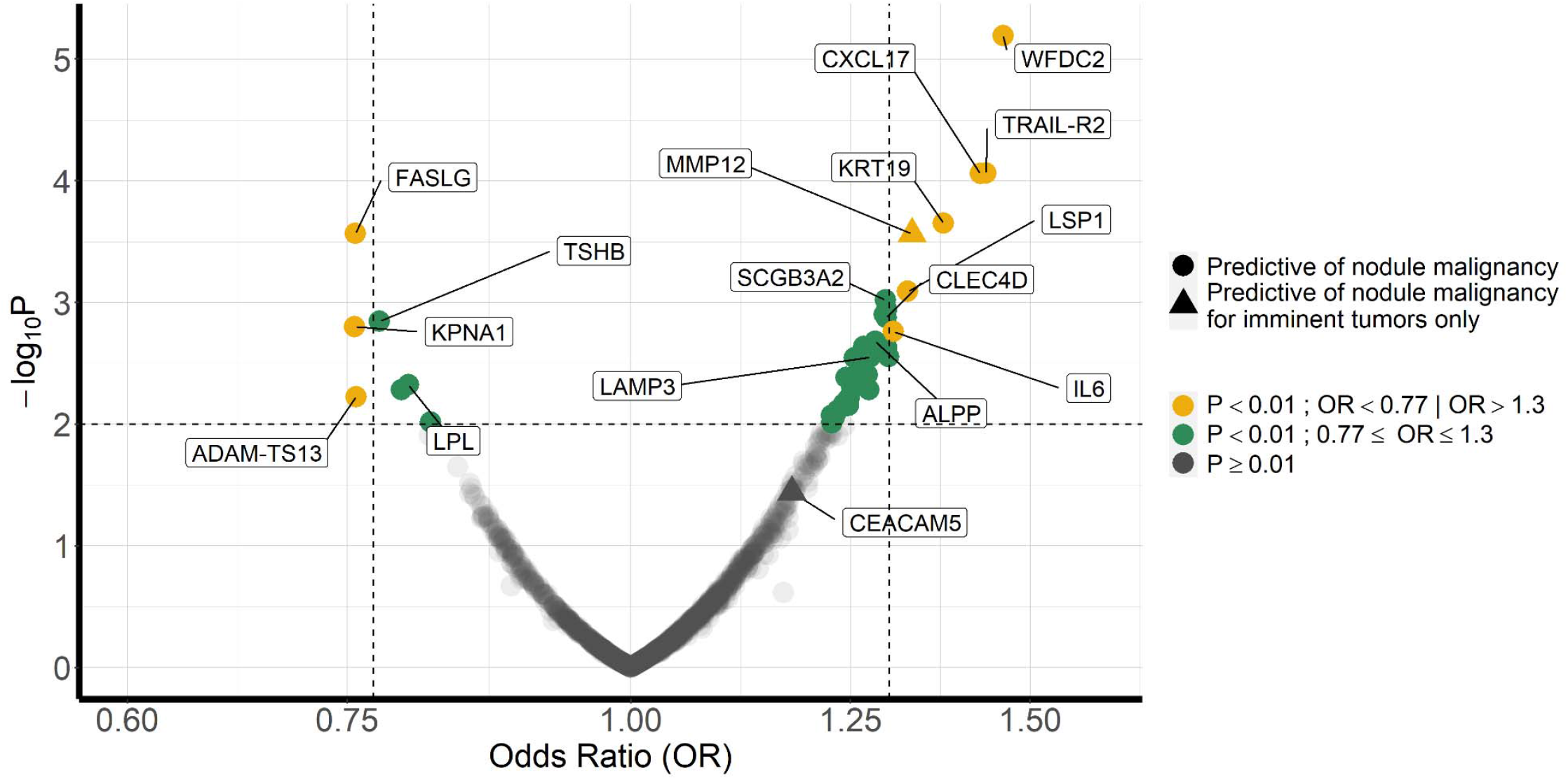
Volcano plot for single-marker analyses of nodule malignancy. The results of individual marker are based on multivariable logistic regression adjusted for Brock Score. Imminent tumor is defined as lung cancer cases diagnosed within one year post blood test.

### Biological relevance of informative protein markers

Based on LASSO and random forest (**Figure 1**), we identified a total of 36 protein markers that were deemed informative of pulmonary nodule malignancy. The full list of the 36 informative markers is shown in the **Supplementary Table 1**. The biological pathways and network that are represented by these 36 potentially informative markers are shown in **Figure 3**. Out of 63 pathways at the levels 2 and 3 of GO-BP, 10 pathways were significantly enriched with FDR less than 0.01 (**Figure 3A**). The most significant pathway is response to external stimulus (FDR<0.001), and the representation of this pathway is approximately 7 times higher than expected by chance when comparing to other pathways. Other pathways that had enriched representation included cell death, immune response, immune system development and more. The network analysis indicated that these 10 pathways are highly interconnected (**Figure 3B**).

**Figure 3:**
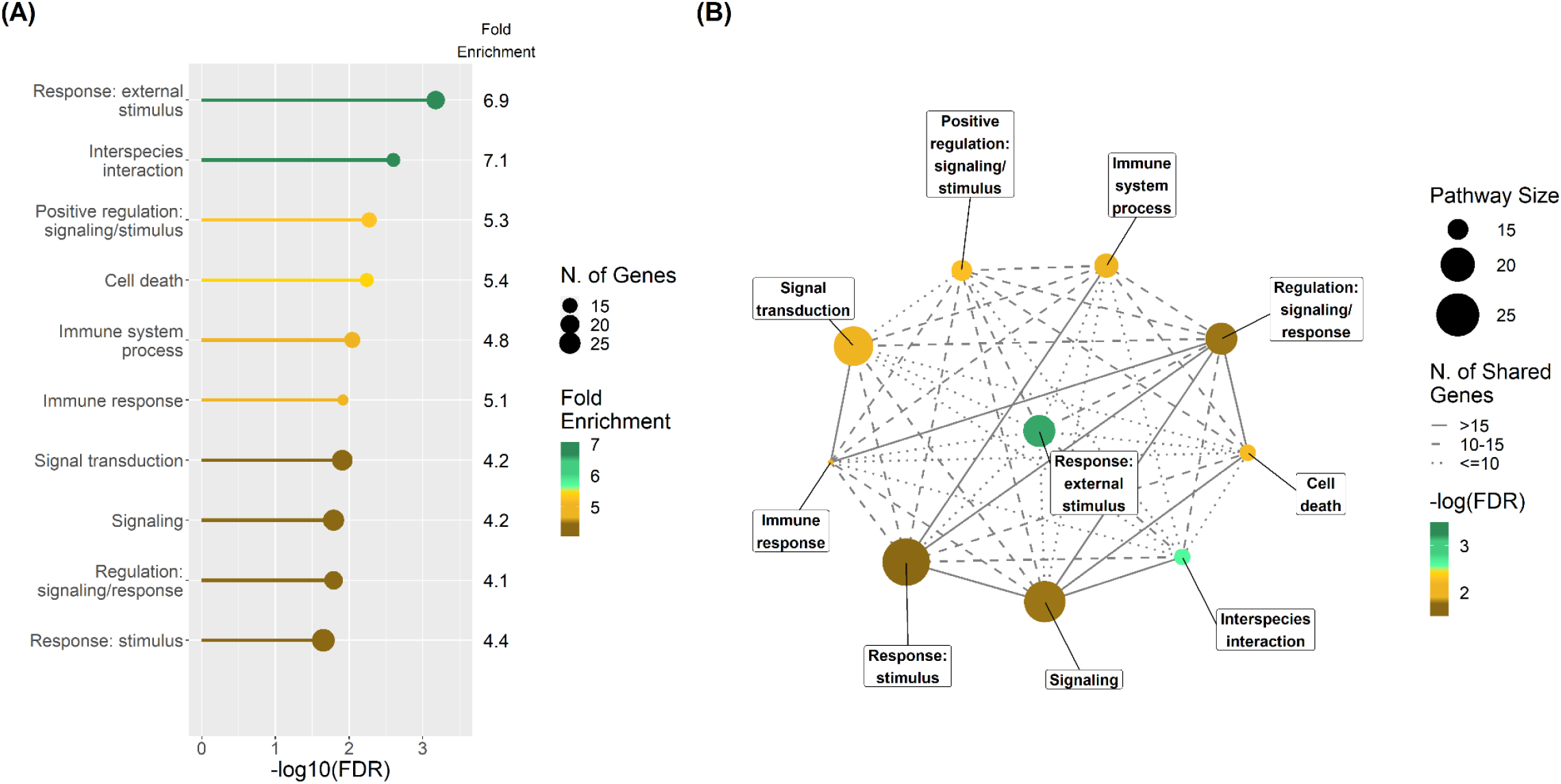
The results of Gene Ontology – Biological Process pathway enrichment analysis and the network analysis based on the 36 informative markers. **(A)** the Bar Plot of the significantly enriched pathways (FDR<0.01) ordered by the FDR levels, with Fold Enrichment value and pathway size (number of genes) annotated. **(B)** The network of the significantly enriched pathways (FDR<0.01), depicting the pathway size (circle), the significance level (gradient), and the connections between pathways (line types).

To understand the role of these top protein markers in cancer hallmarks, we summarized the allocations of our top protein markers to different cancer hallmarks [39] (**Supplementary Figure 2**). Cancer hallmarks include the instrumental milestones for a normal cell to become malignant and survive, proliferate, or spread. Fifteen of the 36 informative circulating protein markers were included in the cancer hallmarks previously described. Among those, FASLG and interleukin 6 (IL6) are involved in over half of those cancer hallmarks. The hallmarks that are most relevant for these circulating protein markers are inducing angiogenesis, resisting cell death and tumor promoting inflammation (**Supplementary Figure 2**).

The unsupervised hierarchical clustering using TCGA mRNA data of the top 35 informative markers (one removed due to data unavailability) showed that mRNA expressions of protein-coding genes have differential profiles by tumor status comparing lung tumor tissues and adjacent normal lung tissues. These differences were not explained by smoking, sex, age of the patients, or tumor characteristics such as location and stage (**Supplementary Figure 3**), and this observation applies to both lung squamous cell carcinoma and adenocarcinoma. This supports the idea that the cancer-associated plasma proteins selected are related to malignant transformations of normal lung tissues.

### Classification performance

From 36 informative markers, we narrowed down to nine markers with Boruta feature selection algorithm after accounting for Brock score based on their discriminatory ability to correctly classify nodule malignancy within 5 years of lead time from blood collection to cancer diagnosis. These markers were WFDC2, FASLG, CXCL17, TRAIL-R2, KRT19, thyroid stimulating hormone subunit beta (TSHB), secretoglobin family 3A member 2 (SCGB3A2), Fc receptor like 5 (FCRL5), and Karyopherin subunit alpha 1 (KPNA1). Based on the nested cross-validation approach, we found that the combination of these nine markers increased the overall sensitivity to 0.80 (p-value<0.001) compared to the Brock score, but did not improve the specificity (p-value=0.135). The area under the receiver operating curve (AUC) from 0.71 with the Brock Score alone to 0.83 (p-value<0.001) (**Figure 4**). When we set the probability threshold to match the performance of LungRADS, the Combined model also outperformed the lungRADs classifications significantly: With the same sensitivity level of 0.59 based on LungRADS category 4, the combined model significantly increased the specificity from 0.79 (LungRADs) to 0.90 (**Table 2**). With the same specificity level of 0.79, the Combined model increased the specificity from 0.59 (LungRADs) to 0.74.

**Figure 4:**
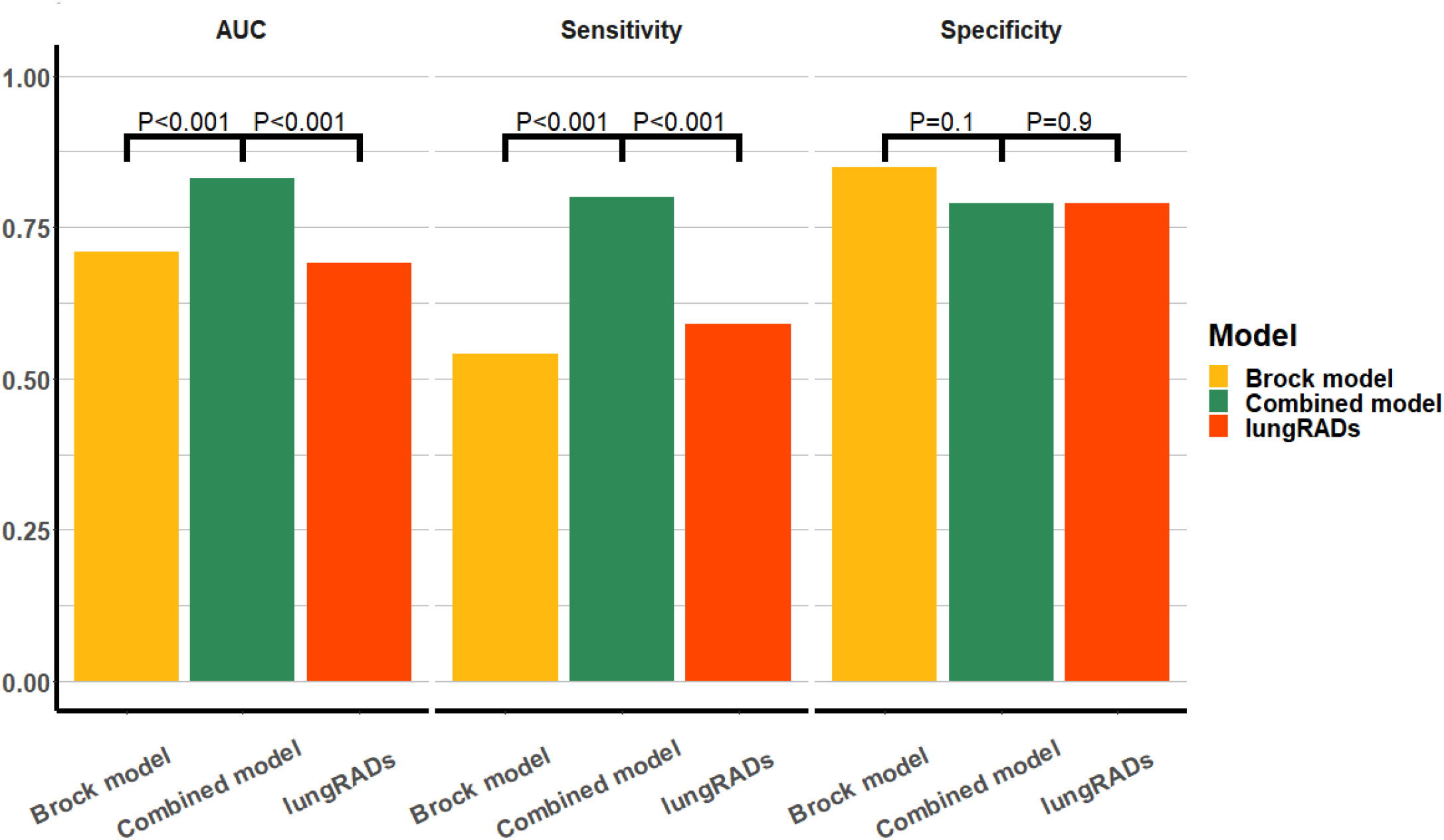
Overall AUC, sensitivity and specificity values of Brock model, LungRADs and Combined model (adding circulating protein markers) for nodule malignancy differentiation. The accuracy measures are calculated based on the threshold corresponding to the largest Youden’s Index. P-values indicates the statistical difference between the corresponding accuracy measures in comparison.

**Table 2:**
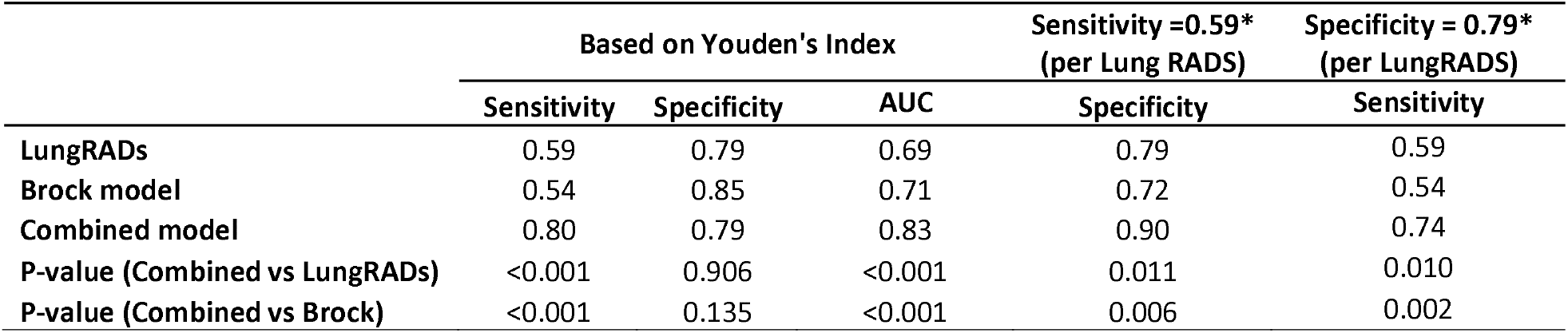
Overall accuracy measures of Brock model, LungRADs and Combined model (adding circulating protein markers) for nodule malignancy differentiation. The accuracy measures are calculated based on the threshold corresponding to the largest Youden’s Index, or by matching the sensitivity and specificity to LungRADs category 4.

### Consideration of key factors

When stratified by smoking status and nodule sizes, the combined model demonstrated better sensitivity compared to Brock score alone in most strata (**Figure 5**), while the specificity remained largely comparable. The only exception is when stratified by nodule sizes, we observed an improvement of specificity by 30% among those with small pulmonary nodules (≤6mm diameter) when adding protein markers (**Figure 5**). This is likely due to the fact that the Brock score is largely driven by nodule size and maybe under-performing for small nodules. In general, both sensitivity and specificity are higher when the time to diagnosis is shorter (**Figure 5**). The AUCs were also improved compared to Brock score alone in all major strata (**Supplementary Figure 4**).

**Figure 5:**
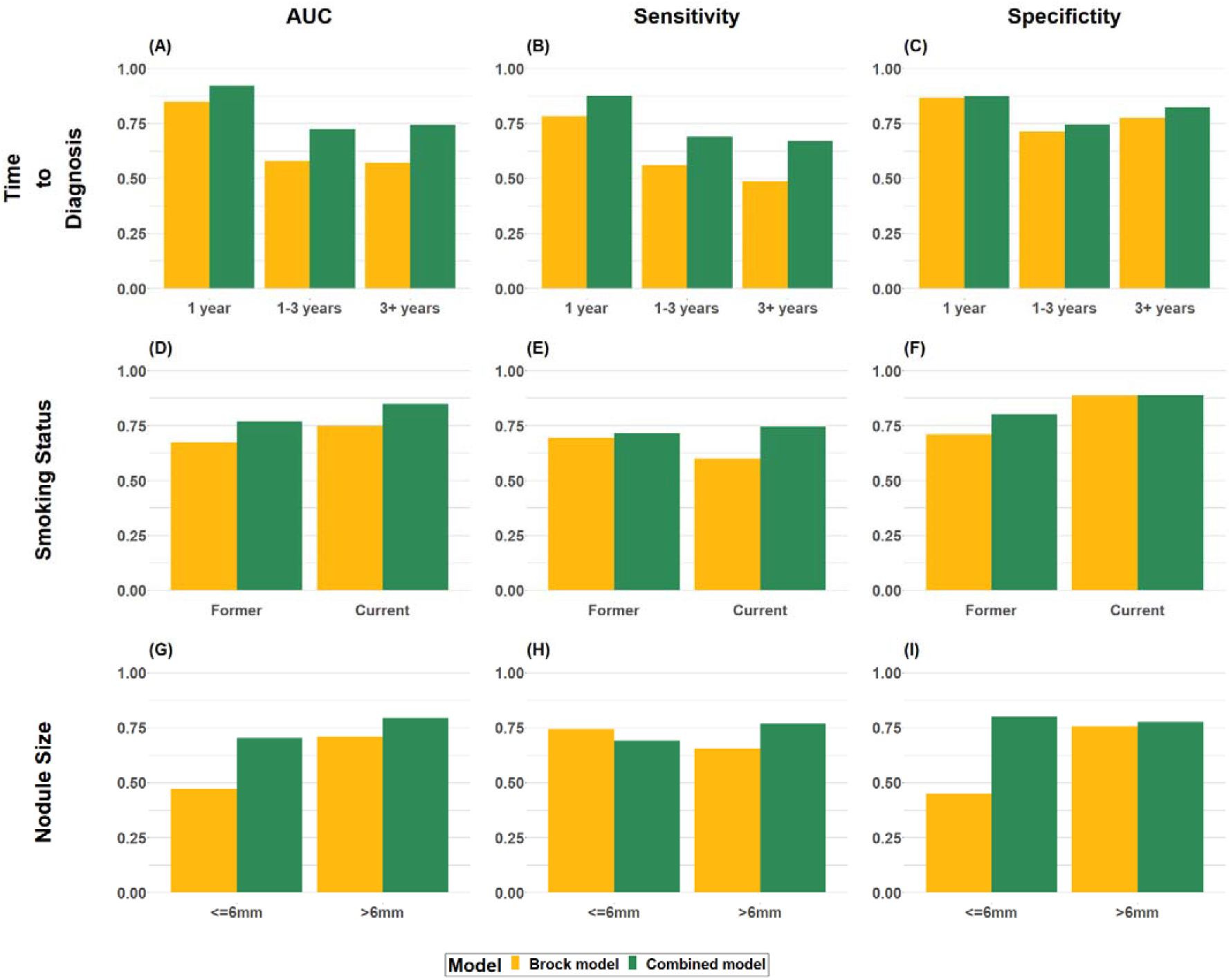
Accuracy measures of Brock model vs. Combined model (Brock score + biomarkers) for nodule malignancy by time to diagnosis (A, B, C), smoking status (D, E, F) and the longest diameter of the nodules (G, H, I).

Specifically for lung cancer diagnosed within 1 year, additional two protein markers were identified, MMP12 and CEACAM5. More information about the 11 protein-coding genes (including the two protein markers that indicate imminent lung cancer) are summarized in **Supplementary Table 2**, including their expression in lung and immune-system related tissues, based on protein and mRNA expression data in the Human Protein Atlas [40, 41] and ProteomicsDB [42, 43]. According to the Human Protein Atlas, all top 11 genes are expressed in lung cancer at transcriptional level. As for protein expression in different anatomical locations, all proteins (when data are available) are observed in lung tissues, except FASLG and TSHB. However, adding the two markers that indicate imminent tumors to the model, the overall classification accuracy did not increase, therefore all the results presented herein are based on 9 markers only.

The distribution of the protein levels for selected markers in healthy individuals, individuals with benign pulmonary nodules and cancer patients are shown in **Supplementary Figure 5**. In general, the distribution of the protein expression in healthy individuals and those with benign nodules were comparable, except for WFDC2 (trend p<0.001) and CEACAM5 (trend p<0.001) where there were notable trends of distribution in the 3 groups. The detailed associations between each of these circulating protein markers and nodule malignancy, by CT screening studies, sex, smoking status, time to diagnosis, histology and nodule size are shown in **Supplementary Figure 1**. We did not observe strong heterogeneity across the 4 LDCT studies for most of the markers. Apart from the notable differences by time to diagnosis for KRT19, MMP12, CEACAM5, we also observed histological differences for SCGB3A2, where no association was observed for squamous cell carcinoma, and by sex for TRAIL-R2, KRT19, and MMP12 where the association was predominately shown among males (**Supplementary Figure 1**).

## DISCUSSION

This study represents the first systematic search for circulating protein markers for pulmonary nodule malignancy. Based on an extensive assessment of circulating proteomics analysis of pre-diagnostic samples from four LDCT screening studies, we showed that the circulating protein markers can help differentiate benign from malignant pulmonary nodules detected in the LDCT screening before clinical cancer diagnosis. The 36 informative markers predominately represent a tightly connected network of pathways related to response to stimulus, cell death and immune system. The circulating protein markers increased the overall classification accuracy and sensitivity, and in the case of small nodules, the specificity was increased.

Given the increasing use of LDCT screening for lung cancer, there has been a growing interest in the recent years in differentiating the malignant and benign pulmonary nodules via the use of the blood-based biomarkers [35, 44-46]. Previous work on biomarkers for nodule malignancy were based on various study designs and target molecules [44, 47], although most had small sample sizes, focused on limited number of markers, and none took a comprehensive and systematic search strategy [48-51]. Given that a single protein marker is unlikely to reach sufficient discriminative power, the advantage of a systematic search we used in the current study allowed us to extract the most informative signals, including novel biomarkers that were not previously investigated.

Some previous studies used pre-diagnostic samples, but none were beyond 2 years prior to diagnosis. We included an expanded time horizons to 5 years before diagnosis, which allowed us to investigate biomarkers by time intervals, and we observed that certain circulating protein markers can differentiate nodule malignancy even more than 3 years before diagnosis (e.g., WFDC2, CXL17, TRAIL-R2, SCGB3A2) and the others (KRT19, CEACAM5 and MMP12) indicated a more mid-term to imminent lung cancers. Being able to assess nodule malignancy more than 3 years prior to clinical diagnosis would help to devise a better patient management strategy, and the ability to distinguish those that might become imminent tumors would also facilitate immediate clinical actions to improve patient’s prognosis. Not surprisingly though, we observed a higher sensitivity and specificity with shorter lead time to diagnosis.

Evaluating biomarkers in a longer pre-diagnostic time window also enabled identification of biological pathways beyond inflammatory-related or cancer-related proteins, which were often the focus of previous studies on imminent lung tumor [42]. Our data show that a tight-knit network of pathways that appear to have a role in differentiating malignant from benign nodules are related to response to stimulus, apoptosis, immune system and regulators of these biological steps.

There are several circulating biomarker tests that are commercially available for indeterminant pulmonary nodules, such as EarlyCDT-lung, and Nodify-XL2 [48, 49, 52, 53]. However, EarlyCDT-Lung, that comprises of 7 autoantibodies, has shown insufficient sensitivity and currently is not recommended by the National Institute for Health and Care Excellence (NICE) guidelines for nodule classification due to weak evidence [54, 55]. Nodify-XL2, comprised of two biomarkers of LG3BP and C163A, has yet to be further validated in the clinical setting [49].

Some of the previous circulating protein panels proposed for indeterminate pulmonary nodules were adapted from the initial discovery for lung cancer occurrence [48]. While the protein expression levels can be comparable for some protein markers, we observed that the expression levels of specific protein markers were different based on the presence of pulmonary nodules, regardless of their subsequent malignancy status, which supports the rationale of using patients with benign nodules as the primary comparison group and healthy controls only to assess background expression level. A notable example is WFDC2, whose expression level increased from healthy, benign nodule to malignant nodules. Using healthy controls without nodules would likely result in over-estimation of the association when the goal is to differentiate benign from malignant nodules.

Some of our 36 informative markers were previously reported to be associated with lung cancer risk, for example interleukin 6 (IL-6), CEA and KRT19, but many were novel markers that have not been previously reported in lung cancer [47, 56, 57]. Fewer were reported for pulmonary nodule malignancy. Two of our top protein markers (CEACAM5 and KRT19) have been used widely in the clinic as general tumor markers, particularly those markers that are associated with imminent tumor in our dataset. For example CEACAM5, a member of human carcinoembryonic antigen (CEA) protein family, is often used to monitor treatment response [58]. Substantial previous work showed that CEACAM5 can be used to predict lung cancer risk, and some evidence suggested the utility of distinguishing between benign and malignant pulmonary nodules [45, 50, 52, 57, 59]. We only observed an association between CEACAM5 and lung cancer within 1 year of lead time. In accordance with our finding regarding the overexpression of CEACAM5 protein in imminent lung cancer tumor, CEACAM5 is known to accelerate the tumor growth and is specifically identified as a metastatic driver [60].

Similarly, KRT19 (or cytokeratin 19) is responsible for structural integrity of epithelial cells, and its fragment antigen (CYFRA21-1) is frequently used to monitor tumor presence, although its specificity for cancer is considered low. It has been previously suggested as a biomarker for both lung cancer risk and nodule malignancy [45, 50-52, 57, 59]. We found that KRT19 is a predictor of nodule malignancy only for the imminent lung tumors, and this is compatible with a previous study showing that KRT19 is released into the blood circulation when necrosis occurs in lung tumor tissue [61].

As one of the most studied multifunctional cytokines, IL-6 is known for its immunosuppressive role in tumorigenesis. It is widely reported that the elevated serum level of cytokine family including IL-6 is associated with lung cancer incidence [62, 63]. However, its role for identifying the indeterminate nodules has not been previously investigated.

Overall, WFDC2 (also known as Human Epididymis Protein 4, HE4) exhibited the strongest association with nodule malignancy in our study. It is known to be expressed in the pulmonary epithelial cells and was previously shown to be associated with innate immunity with a particular role in defense of the lung epithelial cells [41, 64]. Several recent studies have suggested its involvement in COPD and lung cancer through pro-inflammatory responses, and high concentration of HE4 in serum and lung cancer tissues was previously reported [65-67]. It was shown to be associated with lung cancer risk [56], but the associations with nodule malignancy was inconclusive [68].

The remaining of the top markers have not been previously investigated for the differentiation between the malignant and benign nodules; some of which were previously shown to have a role in lung carcinogenesis. As expected, several of the top markers are involved in the inflammation pathway. MMP12, secreted by inflammatory macrophages, was shown to be highly expressed in lung cancers and can trigger angiogenesis and results in inflammatory cell infiltration and epithelial growth [69, 70]. MMP12 was also identified as a tumor-associated antigen (TAA) which triggers immune response and was suggested to be used in the diagnostic autoantibody panel[71, 72]. CXCL17 is a mucosal chemokine that also exhibits angiogenic effect [73] It was shown to be expressed in lung airways and in lung cancer cells [73]. SCGB3A2, a multifunctional secreted protein, can activate inflammasome pathway and lead to pyroptosis [74-76].

Related to programmed cell death, FASLG plays a role in apoptosis signaling and tissue homeostasis, and its loss of expression was often observed in NSCLC [77]. The lower activation of FAS/FASLG signaling pathway implies resisting activation-induced cell death and hence weakens the immune response [78-80]. We observed a lower FASLG level in circulation among lung cancer patients leading to an inverse association, which is consistent with these previous experimental data. TRAIL-R2 was shown to be associated with apoptosis and selectively induce cell death, and it was recently suggested as a negative regulator of the tumor suppressor *p53*, and highly expressed in many types of tumor cells [81]. Given its key role in tumor necrosis factor related apoptosis, several clinical trials have been conducted using TRAIL-R2 agonistic antibodies [82-84]. Involved in evading immune system destruction, FCRL5, a member of immunoglobin receptor family, can inhibit B cell proliferation, which has a critical role in cancer development [85, 86].

In general, the protein markers we identified have differential expression level in lung normal and lung tumor tissues as reported in previous studies. In addition, we observed a distinct mRNA expressions signature based on the genes encoding our 36 informative protein markers for the tumor tissues using TCGA data.

There are several limitations in our study. First, even with a total of 4 LDCT studies, our sample size is modest which may lead to some degree of false-negative findings. To address the issue of statistical instability, we applied a resampling approach, as well as FDR to adjust for multiple comparison. The nested cross-validation approach is considered to be optimal when the sample size is limited [87]. Second, as this study was conducted on a lung cancer screening cohort, the results are not generalizable to lung cancer low-risk cohort including never-smokers. Third, since the NPX values are based on relative abundance, it is not suitable for building a specific algorithm for prediction or classification. Therefore, our main goal was to identify the potential informative markers and the biological pathways that they represent. The assessment of classification accuracy reported within this work is primarily to assess whether there is possible added values for circulating biomarkers compared to nodule characteristics alone. Our next step is to validate the performance of the top markers based on the absolute quantification values, which will allow the construction of predictive algorithm for external validation of an independent study.

There are notable strengths of our study. This represents the first systematic investigation of circulating protein for pulmonary nodule malignancy based on an international collaboration from national screening cohorts. With pre-diagnostic samples, we were able to identify several novel and informative protein markers for pulmonary nodule malignancy. In general, we have observed consistency of results across studies, which suggests the robustness of our results. Our findings are now contributing to the configuration of customized panel for both lung cancer risk and pulmonary nodule malignancy. The findings from our study clearly demonstrated an added value of circulating protein markers when combined with clinico-epidemiological data and the Brock score. This study paves the way to analyze the top markers to be measured in absolute quantifications, which will then permit construction of a probability algorithm and assess model calibration.

In conclusion, we demonstrated that circulating protein markers can help to differentiate malignant from benign pulmonary nodules detected in the LDCT screening. Clinically, this means that the protein markers might be useful to devise a better management plan the patients with smaller nodules with improved specificity, which are often not followed up if there are no other clinical indications. Our study provides a roadmap for developing the protein marker panels for use in pulmonary nodule management after LDCT screening.

## Data Availability

The dataset presented in this study is available from the corresponding author upon request and committee approval.

## Notes

**Funding** This work is supported by NIH U19 CA203654 (INTEGRAL) and Canadian Institute for Health Research (FDN 167273). LMM was also supported by ISCIII Fondo de Investigación Sanitaria-Fondo Europeo de Desarrollo Regional (PI19/00098; PI22/00451), Lung Ambition Alliance and Fundación Roberto Arnal Planelles.

**Conflict of Interests** Authors declared no conflict of interest.

### Competing Interest Statement

The authors have declared no competing interest.

### Funding Statement

This work is supported by NIH U19 CA203654 (INTEGRAL) and Canadian Institute for Health Research (FDN 167273). LMM was also supported by ISCIII Fondo de Investigacióin Sanitaria-Fondo Europeo de Desarrollo Regional (PI19/00098; PI22/00451), Lung Ambition Alliance and Fundación Roberto Arnal Planelles.

### Author Declarations

Ethics committee/IRB of the Mount Sinai Hospital gave ethical approval for this work. The Ethics approval was also obtained from local institutes at each study site.

### Summary of Updates

The manuscript sections are reordered for clarity.

