## Supplementary materials for "Circulating Proteome for Pulmonary Nodule Malignancy"

### Supplementary methods: Description of the LDCT screening cohorts participating in this study

#### PanCan: Pan-Canadian Early Detection of Lung Cancer Study

This study prospectively recruited 2537 subjects between September 2008, and December, 2010 [1]. Participants had at least a 2% 6-year risk of lung cancer as estimated by the PanCan model, a precursor to the validated PLCOm2012 model. The eligible subjects were ever smokers, aged 50-75 years who had no previous history of invasive cancer. The subjects were recruited at eight centers across Canada (Vancouver, Calgary, Hamilton, Toronto, Ottawa, Quebec, Halifax, and St. John's). All study participants completed a detailed questionnaire, spirometry, collection of blood specimens for biomarker measurement and LDCT at baseline. Participants with abnormal LDCT results (the presence of any non-calcified non-perifissural pulmonary nodule or a nodule with non-solid area of at least 1mm diameter) were followed up every 3-24 months. A nodule was considered benign if it showed a benign calcification pattern (e.g., fully calcified, popcorn calcification) or if the size and density of was unchanged for longer than 2 years.

#### UKLS: The UK Lung Cancer Pilot Screening Trial

The UKLS is a randomized controlled trial of LDCT screening in individuals aged 50-75 years with at least 4.5% risk of developing lung cancer in 5 years based on the Liverpool Lung Project (LLPv2) risk model [2, 3]. The subjects with any comorbidity that unequivocally contraindicated either screening or treatment were excluded. There were 4055 individuals recruited in two geographical areas, covering six primary care trusts around Liverpool and Cambridge, from 2011 to 2013. The participants were randomized to receive either LDCT or usual care, 1994 of which underwent LDCT scans. Most participants had their lung function measured, provided blood samples, and completed detailed epidemiological, psychosocial and health economics questionnaires at baseline. The LDCT subjects received a baseline scan and nodules were assessed by two local radiologists and placed in one of four pre-defined nodule categories: Category 4 (Solid: volume >500mm$^3$ or diameter >10mm; Part solid: solid component- volume >500mm$^3$); Category 3 (Solid: volume 50 - 500mm$^3$ or diameter 5 - 9.9mm; Part solid: non-solid component diameter >5mm, solid component :15 - 500mm$^3$ or diameter 3 - 9.9mm; Non-solid: diameter ≥ 5mm), Category 2 (Solid: volume 15 - 49mm$^3$ or diameter 3 -4.9mm; Part solid: solid component volume <15mm$^3$ or diameter < 3mm; Non solid diameter 3 - 4.9mm); Category 1 (Benign nodule or volume <15mm$^3$ or diameter <3mm or features of an intrapulmonary lymph node). Consensus nodule category was assigned following central reading at the Royal Brompton Hospital, with a read for arbitration when necessary. Category 4 nodules were immediately referred for work-up and clinical management. Category 3 nodules identified in the baseline scan were re-analyzed in follow-up CT scans at three and twelve months; Category 2 nodules at twelve months only. Category 1 nodules were considered normal and had no further follow-up. Growth of nodules was based on their characteristics and volume doubling time (VDT); i.e. VDT <400 days or new solid component of non-solid nodule was classified as growth in the UKLS trial and were referred to the trial participating MDT clinic for work-up and clinical management.

**IELCAP-Toronto: International Early Lung Cancer Action Program – Toronto**

This single arm early detection of lung cancer research study at Princess Margaret Cancer Centre, Toronto, Canada joined the international Early Lung Cancer Action Program (I-ELCAP) in 2003 [4, 5]. Ever smokers of more than 10 pack-years, aged 50 years or older were eligible for LDCT screening. A total of 4782 individuals have been enrolled in the Greater Toronto Area, and 4161 of those have the clinic-epidemiological data needed to be part of INTEGRAL. Participants were administered a LDCT scan along with a standard study questionnaire at baseline. Blood samples were systematically collected at baseline since 2006. In this study, a LDCT results was considered to be normal if no nodules of the size of 5 mm or larger was found. In case of a positive LDCT result, an indeterminate noncalcified solid or part-solid nodule 5 mm or larger, or a nonsolid nodule 8 mm or larger was detected. The abnormal scans were followed up at 1-2 years depending on nodule size and shape. The location, size, consistency, calcifications and spiculations were determined for each nodule.

**P-IELCAP: Pamplona International Early Lung Cancer Action Program**

P-IELCAP is an ongoing lung cancer screening program in Spain which began recruitment in September 2000 [6]. It is part of the I-ELCAP consortium. As of December 2019, the program had enrolled 3825 asymptomatic subjects older than 40 years of age, including former of current smokers (≥10 pack-years), cancer-free 5 years prior to enrollment. The subjects completed LDCT, spirometry, blood collection and a health questionnaire at baseline and during follow up. Subjects with a positive baseline LDCT (noncalcified nodules ≥5 or 6 mm depending on the year of enrollment), had imaging follow up at 1-3 months. Subjects with negative scans or small pulmonary nodules at baseline had follow up LDCTs at annual intervals.

**Supplementary Table 2: Summary of the top 11 informative circulating protein markers**

| Gene name | Full name (Ensembl Gene ID) | Cytoband | Expression in selected tissues focuses on lung and immune system | | | |
|---|---|---|---|---|---|---|
| | | | Normal tissues: Human Protein Atlas (HPA) [7]and Proteomics DB (PDB) [8-10] | | Cancer tissues: Human Protein Atlas (HPA)[7] | |
| | | | Protein expression | mRNA expression-HPA | Protein | mRNA |
| WFDC2 | WAP four-disulfide core domain 2 (ENSG00000101443) | 20q13 | **HPA:** Bronchus$^{H}$<br>**PDB:** Lung | Lung | Lung cancer | Lung cancer |
| FASLG | Fas ligand (ENSG00000117560) | 1q24 | **HPA:** Bone marrow$^{M}$, Lymph node$^{L}$<br>**PDB:** Data not available | Lymph node, Lung | No expression | Lung cancer |
| CXCL17 | C-X-C motif chemokine ligand 17 (ENSG00000189377) | 19q13 | **HPA:** Data not available<br>**PDB:** Lung | Lung | Data not available | Lung cancer |
| TSHB | Thyroid stimulating hormone subunit beta (ENSG00000134200) | 1q13 | **HPA:** None<br>**PDB:** None | Lung | No expression | Lung cancer |
| TRAIL-R2 (TNFRSF10B) | TNF receptor superfamily member 10b (ENSG00000120889) | 8p21 | **HPA:** Bronchus$^{H}$, Lung$^{H}$, Lymph node$^{M}$<br>**PDB**: Lung, Blood platelet | Lung, Bone marrow, Lymph node | Lung cancer | Lung cancer, Blood platelet |
| SCGB3A2 | Secretoglobin family 3a member 2 (ENSG00000164265) | 5q32 | **HPA:** Data not available<br>**PDB:** Lung | Lung | Data not available | Lung cancer |
| FCRL5 | Fc receptor like 5 (ENSG00000143297) | 1q23 | **HPA:** Data not available<br>**PDB:** Lung | Lymph node, Bone marrow, Lung | Data not available | Lung cancer |

| | | | | | | |
|---|---|---|---|---|---|---|
| KRT19 | Keratin 19 (ENSG00000171345) | 17q21 | **HPA:** Bronchus$^{H}$, Lung$^{H}$ **PDB:** Lung, Lymph node, Cytotoxic T-lymphocyte, Blood platelet | Lung | Lung cancer | Lung cancer |
| KPNA1 | Karyopherin subunit alpha 1 (ENSG00000114030) | 3q21 | **HPA:** Lung$^{L}$, Lymph node$^{L}$ **PDB:** Natural killer cell, B-lymphocyte, Cytotoxic T-lymphocyte, Monocyte, Lymph node, Lung, Helper T-lymphocyte, Cardia, Blood platelet | Lymph node, Lung | Lung cancer | Lung cancer |
| MMP12 | Matrix metallopeptidase 12 (ENSG00000262406) | 11q22 | **HPA:** Data not available **PDB:** Lymph node, Lung, Bone marrow, Blood platelet | Lymph node, Lung | Data not available | Lung cancer |
| CEACAM5 | Cea cell adhesion molecule 5 (ENSG00000105388) | 19q13 | **HPA:** Lung$^{L}$, Bone marrow$^{L}$ **PDB:** Lung | Lung | Lung cancer | Lung cancer |

The Human Protein Atlas (HPA); H: High; M: Medium; L: Low
ProteomicsDB (PDB)

**Supplementary Figure 1: Forest plots of the top circulating protein markers illustrating the association with nodule malignancy stratified by study site, time to diagnosis, smoking status, sex, histology and nodule size, after adjustment for Brock score. The Odds Ratios (OR) and the 95% Confidence Intervals (CI) are presented.**

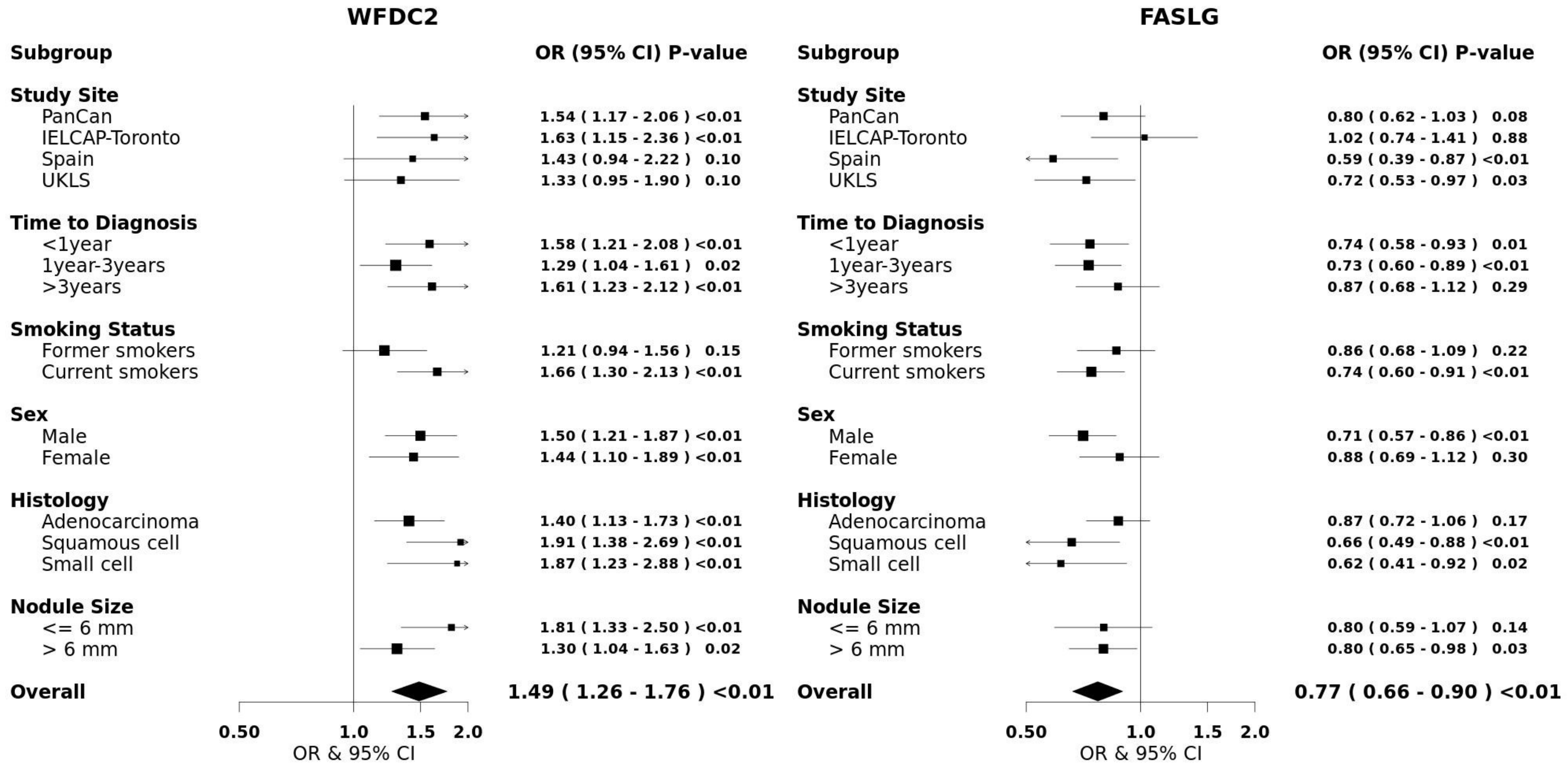

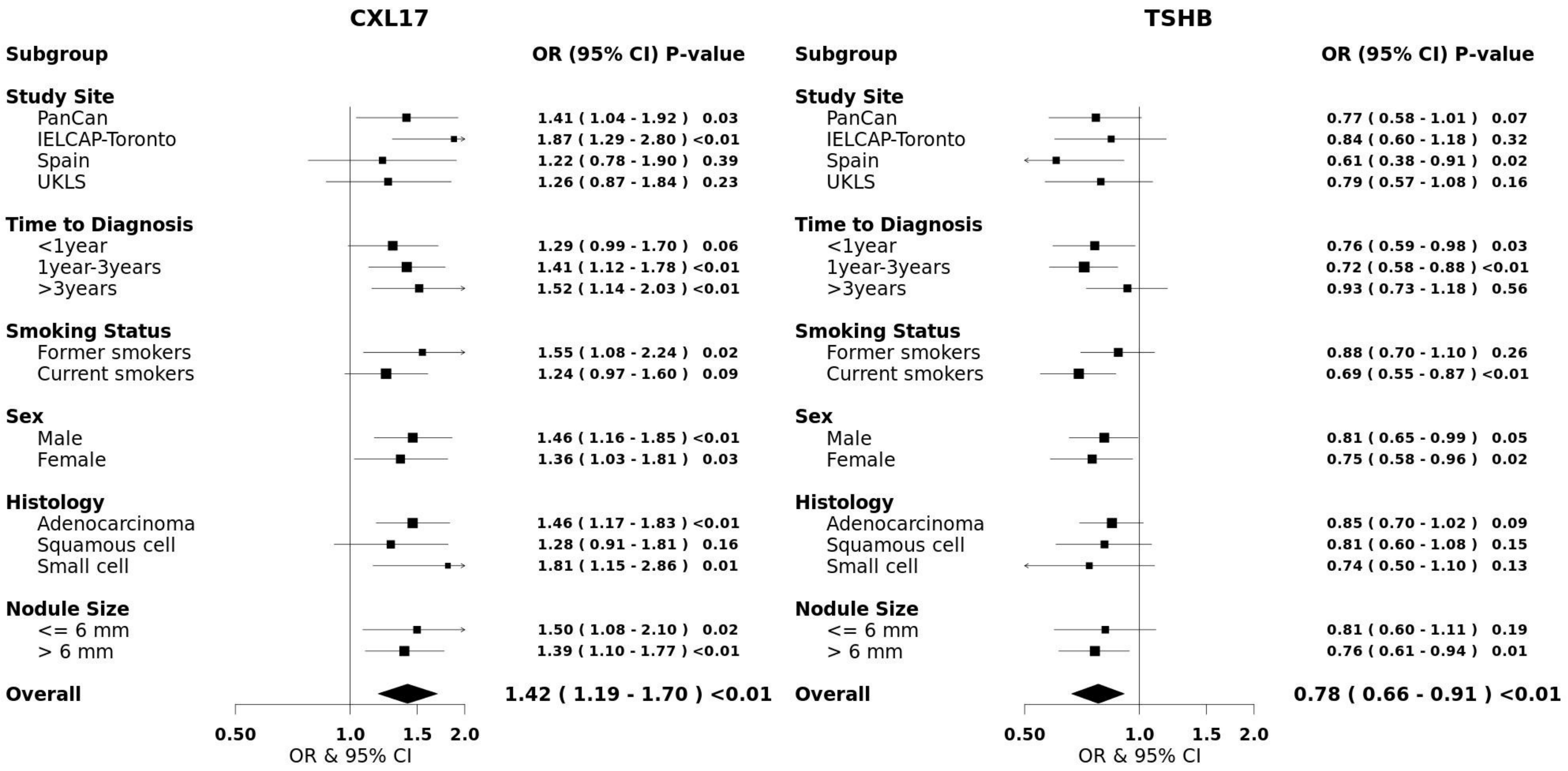
CXL17
Subgroup
OR (95% CI) P-value
Study Site
PanCan 1.41 ( 1.04 - 1.92 ) 0.03
IELCAP-Toronto 1.87 ( 1.29 - 2.80 ) <0.01
Spain 1.22 ( 0.78 - 1.90 ) 0.39
UKLS 1.26 ( 0.87 - 1.84 ) 0.23
Time to Diagnosis
<1year 1.29 ( 0.99 - 1.70 ) 0.06
1year-3years 1.41 ( 1.12 - 1.78 ) <0.01
>3years 1.52 ( 1.14 - 2.03 ) <0.01
Smoking Status
Former smokers 1.55 ( 1.08 - 2.24 ) 0.02
Current smokers 1.24 ( 0.97 - 1.60 ) 0.09
Sex
Male 1.46 ( 1.16 - 1.85 ) <0.01
Female 1.36 ( 1.03 - 1.81 ) 0.03
Histology
Adenocarcinoma 1.46 ( 1.17 - 1.83 ) <0.01
Squamous cell 1.28 ( 0.91 - 1.81 ) 0.16
Small cell 1.81 ( 1.15 - 2.86 ) 0.01
Nodule Size
<= 6 mm 1.50 ( 1.08 - 2.10 ) 0.02
> 6 mm 1.39 ( 1.10 - 1.77 ) <0.01
Overall 1.42 ( 1.19 - 1.70 ) <0.01
0.50 1.0 1.5 2.0
OR & 95% CI
TSHB
Subgroup
OR (95% CI) P-value
Study Site
PanCan 0.77 ( 0.58 - 1.01 ) 0.07
IELCAP-Toronto 0.84 ( 0.60 - 1.18 ) 0.32
Spain 0.61 ( 0.38 - 0.91 ) 0.02
UKLS 0.79 ( 0.57 - 1.08 ) 0.16
Time to Diagnosis
<1year 0.76 ( 0.59 - 0.98 ) 0.03
1year-3years 0.72 ( 0.58 - 0.88 ) <0.01
>3years 0.93 ( 0.73 - 1.18 ) 0.56
Smoking Status
Former smokers 0.88 ( 0.70 - 1.10 ) 0.26
Current smokers 0.69 ( 0.55 - 0.87 ) <0.01
Sex
Male 0.81 ( 0.65 - 0.99 ) 0.05
Female 0.75 ( 0.58 - 0.96 ) 0.02
Histology
Adenocarcinoma 0.85 ( 0.70 - 1.02 ) 0.09
Squamous cell 0.81 ( 0.60 - 1.08 ) 0.15
Small cell 0.74 ( 0.50 - 1.10 ) 0.13
Nodule Size
<= 6 mm 0.81 ( 0.60 - 1.11 ) 0.19
> 6 mm 0.76 ( 0.61 - 0.94 ) 0.01
Overall 0.78 ( 0.66 - 0.91 ) <0.01
0.50 1.0 1.5 2.0
OR & 95% CI

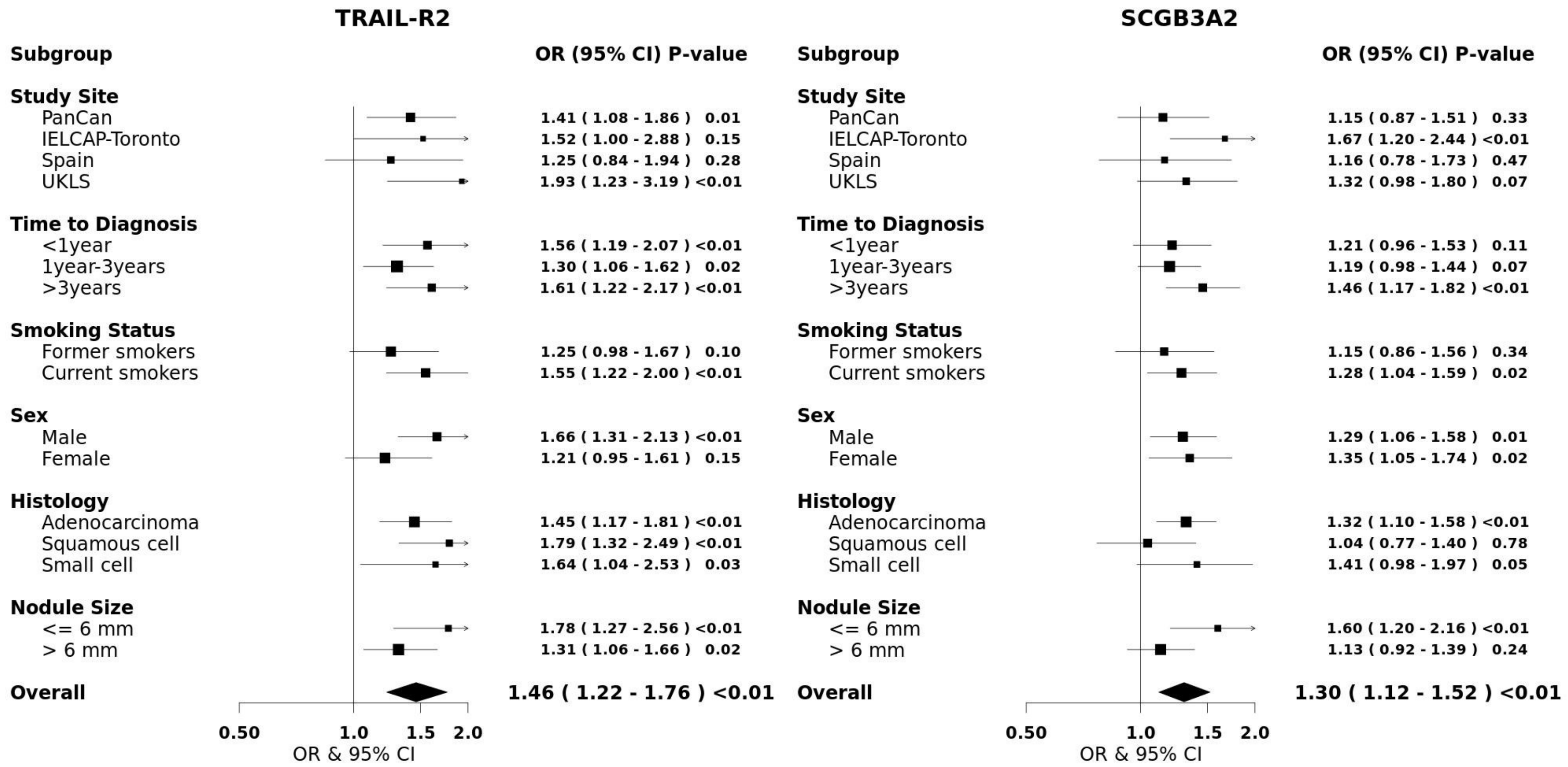
TRAIL-R2
Subgroup
OR (95% CI) P-value
Study Site
PanCan 1.41 ( 1.08 - 1.86 ) 0.01
IELCAP-Toronto 1.52 ( 1.00 - 2.88 ) 0.15
Spain 1.25 ( 0.84 - 1.94 ) 0.28
UKLS 1.93 ( 1.23 - 3.19 ) <0.01
Time to Diagnosis
<1year 1.56 ( 1.19 - 2.07 ) <0.01
1year-3years 1.30 ( 1.06 - 1.62 ) 0.02
>3years 1.61 ( 1.22 - 2.17 ) <0.01
Smoking Status
Former smokers 1.25 ( 0.98 - 1.67 ) 0.10
Current smokers 1.55 ( 1.22 - 2.00 ) <0.01
Sex
Male 1.66 ( 1.31 - 2.13 ) <0.01
Female 1.21 ( 0.95 - 1.61 ) 0.15
Histology
Adenocarcinoma 1.45 ( 1.17 - 1.81 ) <0.01
Squamous cell 1.79 ( 1.32 - 2.49 ) <0.01
Small cell 1.64 ( 1.04 - 2.53 ) 0.03
Nodule Size
<= 6 mm 1.78 ( 1.27 - 2.56 ) <0.01
> 6 mm 1.31 ( 1.06 - 1.66 ) 0.02
Overall 1.46 ( 1.22 - 1.76 ) <0.01
0.50
1.0
1.5
2.0
OR & 95% CI
SCGB3A2
Subgroup
OR (95% CI) P-value
Study Site
PanCan 1.15 ( 0.87 - 1.51 ) 0.33
IELCAP-Toronto 1.67 ( 1.20 - 2.44 ) <0.01
Spain 1.16 ( 0.78 - 1.73 ) 0.47
UKLS 1.32 ( 0.98 - 1.80 ) 0.07
Time to Diagnosis
<1year 1.21 ( 0.96 - 1.53 ) 0.11
1year-3years 1.19 ( 0.98 - 1.44 ) 0.07
>3years 1.46 ( 1.17 - 1.82 ) <0.01
Smoking Status
Former smokers 1.15 ( 0.86 - 1.56 ) 0.34
Current smokers 1.28 ( 1.04 - 1.59 ) 0.02
Sex
Male 1.29 ( 1.06 - 1.58 ) 0.01
Female 1.35 ( 1.05 - 1.74 ) 0.02
Histology
Adenocarcinoma 1.32 ( 1.10 - 1.58 ) <0.01
Squamous cell 1.04 ( 0.77 - 1.40 ) 0.78
Small cell 1.41 ( 0.98 - 1.97 ) 0.05
Nodule Size
<= 6 mm 1.60 ( 1.20 - 2.16 ) <0.01
> 6 mm 1.13 ( 0.92 - 1.39 ) 0.24
Overall 1.30 ( 1.12 - 1.52 ) <0.01
0.50
1.0
1.5
2.0
OR & 95% CI

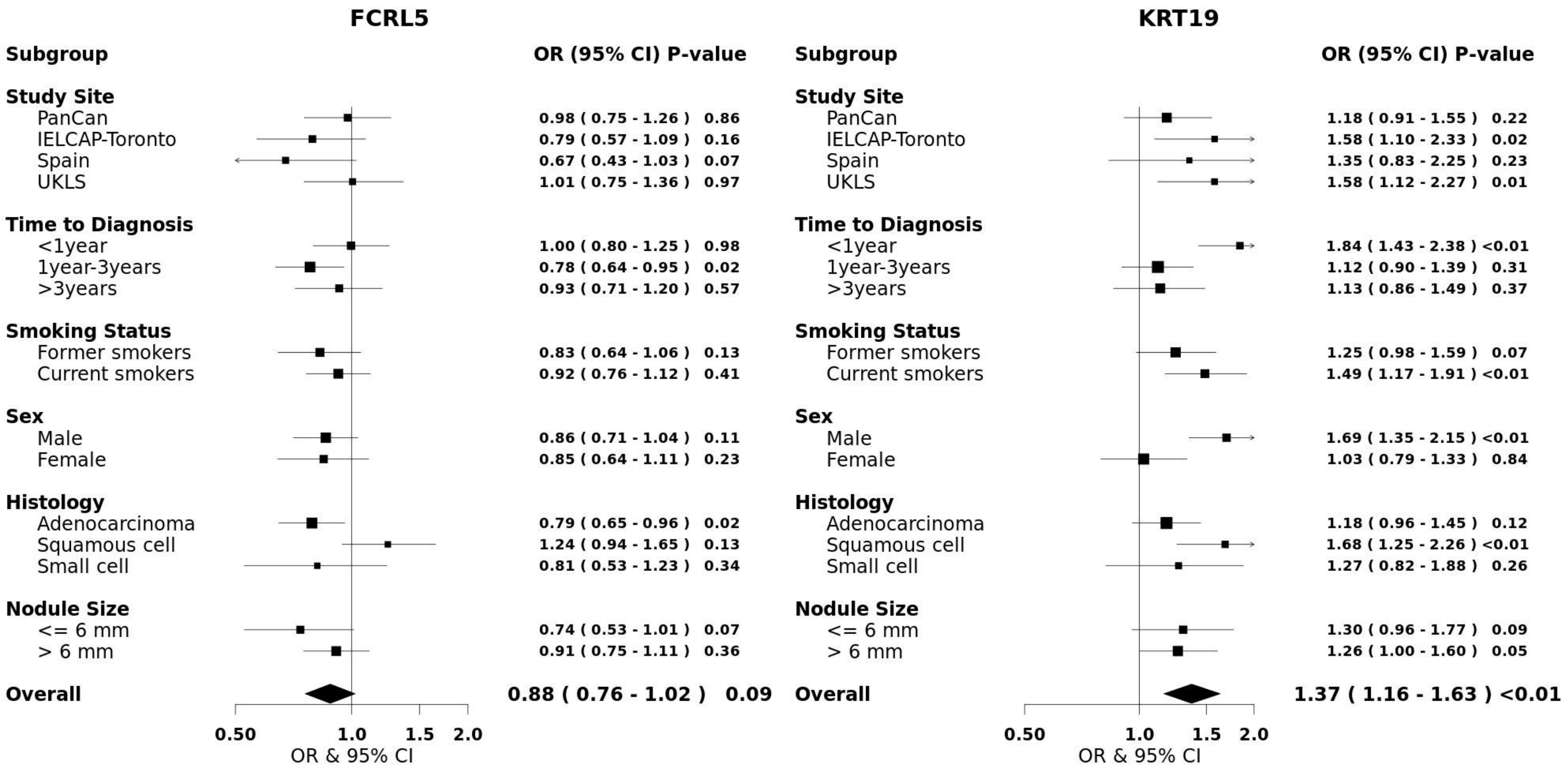
FCRL5
Subgroup
OR (95% CI) P-value
Study Site
PanCan 0.98 ( 0.75 - 1.26 ) 0.86
IELCAP-Toronto 0.79 ( 0.57 - 1.09 ) 0.16
Spain 0.67 ( 0.43 - 1.03 ) 0.07
UKLS 1.01 ( 0.75 - 1.36 ) 0.97
Time to Diagnosis
<1year 1.00 ( 0.80 - 1.25 ) 0.98
1year-3years 0.78 ( 0.64 - 0.95 ) 0.02
>3years 0.93 ( 0.71 - 1.20 ) 0.57
Smoking Status
Former smokers 0.83 ( 0.64 - 1.06 ) 0.13
Current smokers 0.92 ( 0.76 - 1.12 ) 0.41
Sex
Male 0.86 ( 0.71 - 1.04 ) 0.11
Female 0.85 ( 0.64 - 1.11 ) 0.23
Histology
Adenocarcinoma 0.79 ( 0.65 - 0.96 ) 0.02
Squamous cell 1.24 ( 0.94 - 1.65 ) 0.13
Small cell 0.81 ( 0.53 - 1.23 ) 0.34
Nodule Size
<= 6 mm 0.74 ( 0.53 - 1.01 ) 0.07
> 6 mm 0.91 ( 0.75 - 1.11 ) 0.36
Overall 0.88 ( 0.76 - 1.02 ) 0.09
0.50
1.0
1.5
2.0
OR & 95% CI
KRT19
Subgroup
OR (95% CI) P-value
Study Site
PanCan 1.18 ( 0.91 - 1.55 ) 0.22
IELCAP-Toronto 1.58 ( 1.10 - 2.33 ) 0.02
Spain 1.35 ( 0.83 - 2.25 ) 0.23
UKLS 1.58 ( 1.12 - 2.27 ) 0.01
Time to Diagnosis
<1year 1.84 ( 1.43 - 2.38 ) <0.01
1year-3years 1.12 ( 0.90 - 1.39 ) 0.31
>3years 1.13 ( 0.86 - 1.49 ) 0.37
Smoking Status
Former smokers 1.25 ( 0.98 - 1.59 ) 0.07
Current smokers 1.49 ( 1.17 - 1.91 ) <0.01
Sex
Male 1.69 ( 1.35 - 2.15 ) <0.01
Female 1.03 ( 0.79 - 1.33 ) 0.84
Histology
Adenocarcinoma 1.18 ( 0.96 - 1.45 ) 0.12
Squamous cell 1.68 ( 1.25 - 2.26 ) <0.01
Small cell 1.27 ( 0.82 - 1.88 ) 0.26
Nodule Size
<= 6 mm 1.30 ( 0.96 - 1.77 ) 0.09
> 6 mm 1.26 ( 1.00 - 1.60 ) 0.05
Overall 1.37 ( 1.16 - 1.63 ) <0.01
0.50
1.0
1.5
2.0
OR & 95% CI

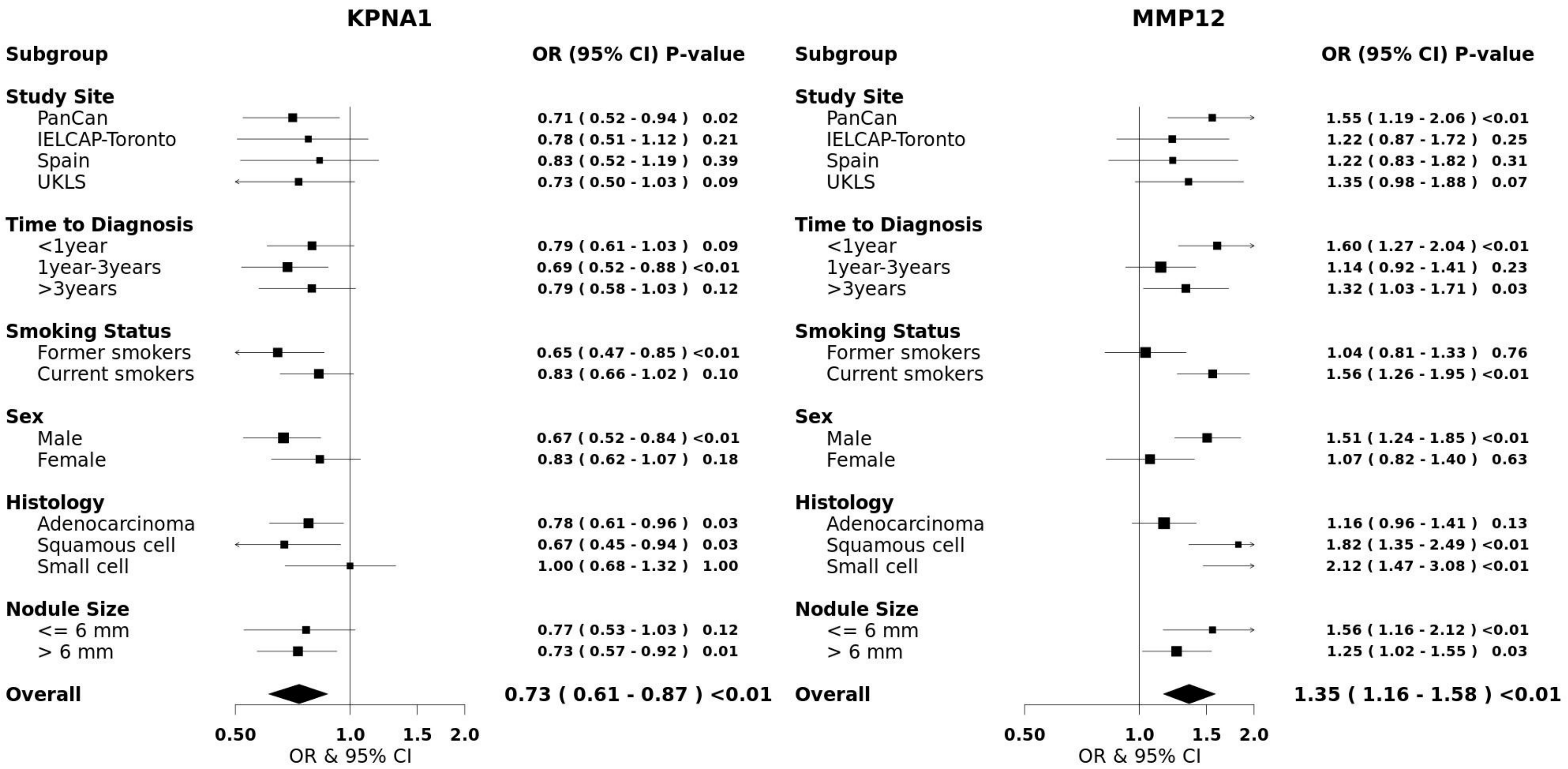
KPNA1
Subgroup
OR (95% CI) P-value
Study Site
PanCan 0.71 ( 0.52 - 0.94 ) 0.02
IELCAP-Toronto 0.78 ( 0.51 - 1.12 ) 0.21
Spain 0.83 ( 0.52 - 1.19 ) 0.39
UKLS 0.73 ( 0.50 - 1.03 ) 0.09
Time to Diagnosis
<1year 0.79 ( 0.61 - 1.03 ) 0.09
1year-3years 0.69 ( 0.52 - 0.88 ) <0.01
>3years 0.79 ( 0.58 - 1.03 ) 0.12
Smoking Status
Former smokers 0.65 ( 0.47 - 0.85 ) <0.01
Current smokers 0.83 ( 0.66 - 1.02 ) 0.10
Sex
Male 0.67 ( 0.52 - 0.84 ) <0.01
Female 0.83 ( 0.62 - 1.07 ) 0.18
Histology
Adenocarcinoma 0.78 ( 0.61 - 0.96 ) 0.03
Squamous cell 0.67 ( 0.45 - 0.94 ) 0.03
Small cell 1.00 ( 0.68 - 1.32 ) 1.00
Nodule Size
<= 6 mm 0.77 ( 0.53 - 1.03 ) 0.12
> 6 mm 0.73 ( 0.57 - 0.92 ) 0.01
Overall 0.73 ( 0.61 - 0.87 ) <0.01
0.50 1.0 1.5 2.0
OR & 95% CI
MMP12
Subgroup
OR (95% CI) P-value
Study Site
PanCan 1.55 ( 1.19 - 2.06 ) <0.01
IELCAP-Toronto 1.22 ( 0.87 - 1.72 ) 0.25
Spain 1.22 ( 0.83 - 1.82 ) 0.31
UKLS 1.35 ( 0.98 - 1.88 ) 0.07
Time to Diagnosis
<1year 1.60 ( 1.27 - 2.04 ) <0.01
1year-3years 1.14 ( 0.92 - 1.41 ) 0.23
>3years 1.32 ( 1.03 - 1.71 ) 0.03
Smoking Status
Former smokers 1.04 ( 0.81 - 1.33 ) 0.76
Current smokers 1.56 ( 1.26 - 1.95 ) <0.01
Sex
Male 1.51 ( 1.24 - 1.85 ) <0.01
Female 1.07 ( 0.82 - 1.40 ) 0.63
Histology
Adenocarcinoma 1.16 ( 0.96 - 1.41 ) 0.13
Squamous cell 1.82 ( 1.35 - 2.49 ) <0.01
Small cell 2.12 ( 1.47 - 3.08 ) <0.01
Nodule Size
<= 6 mm 1.56 ( 1.16 - 2.12 ) <0.01
> 6 mm 1.25 ( 1.02 - 1.55 ) 0.03
Overall 1.35 ( 1.16 - 1.58 ) <0.01
0.50 1.0 1.5 2.0
OR & 95% CI

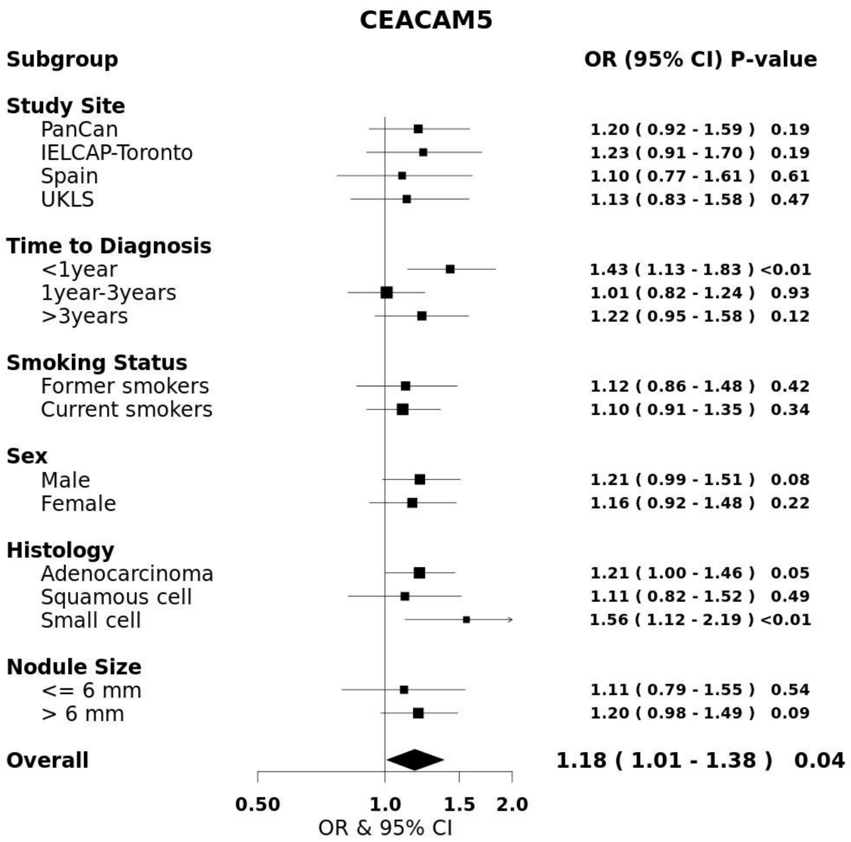
CEACAM5
Subgroup
OR (95% CI) P-value
Study Site
PanCan
1.20 ( 0.92 - 1.59 ) 0.19
IELCAP-Toronto
1.23 ( 0.91 - 1.70 ) 0.19
Spain
1.10 ( 0.77 - 1.61 ) 0.61
UKLS
1.13 ( 0.83 - 1.58 ) 0.47
Time to Diagnosis
<1year
1.43 ( 1.13 - 1.83 ) <0.01
1year-3years
1.01 ( 0.82 - 1.24 ) 0.93
>3years
1.22 ( 0.95 - 1.58 ) 0.12
Smoking Status
Former smokers
1.12 ( 0.86 - 1.48 ) 0.42
Current smokers
1.10 ( 0.91 - 1.35 ) 0.34
Sex
Male
1.21 ( 0.99 - 1.51 ) 0.08
Female
1.16 ( 0.92 - 1.48 ) 0.22
Histology
Adenocarcinoma
1.21 ( 1.00 - 1.46 ) 0.05
Squamous cell
1.11 ( 0.82 - 1.52 ) 0.49
Small cell
1.56 ( 1.12 - 2.19 ) <0.01
Nodule Size
<= 6 mm
1.11 ( 0.79 - 1.55 ) 0.54
> 6 mm
1.20 ( 0.98 - 1.49 ) 0.09
Overall
1.18 ( 1.01 - 1.38 ) 0.04
0.50
1.0
1.5
2.0
OR & 95% CI

**Supplementary Figure 2: The cancer hallmarks represented by the 36 informative protein markers, and the corresponding allocation.** The fold enrichment is calculated for each hallmark separately, depicted by circle size, and the significance level (-logFDR) is annotated by color gradient.

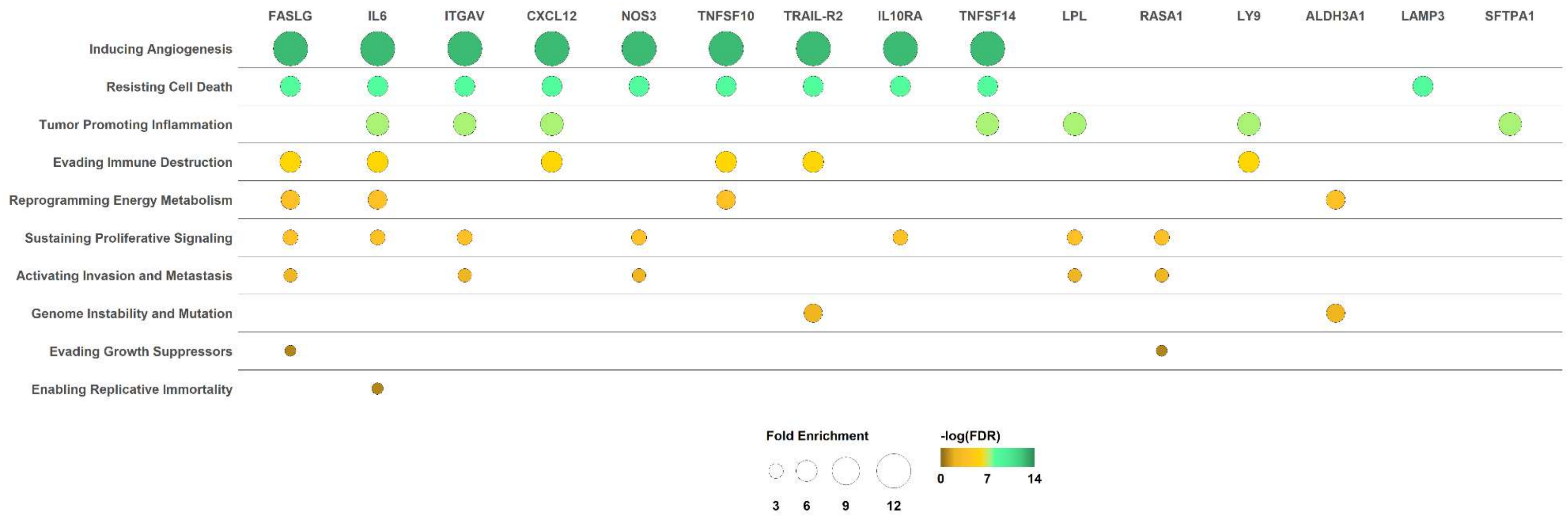

**Supplementary Figure 3: Heatmap of differential gene expressions based on TCGA data comparing lung cancer tissues vs lung normal tissues based on TCGA database by unsupervised hierarchical clustering. (A)** Adenocarcinoma **(B)** Squamous cell carcinoma. This plot includes 35 genes, as TSHB data was not available in TCGA.

**(A) Adenocarcinoma lung cancer tissues**

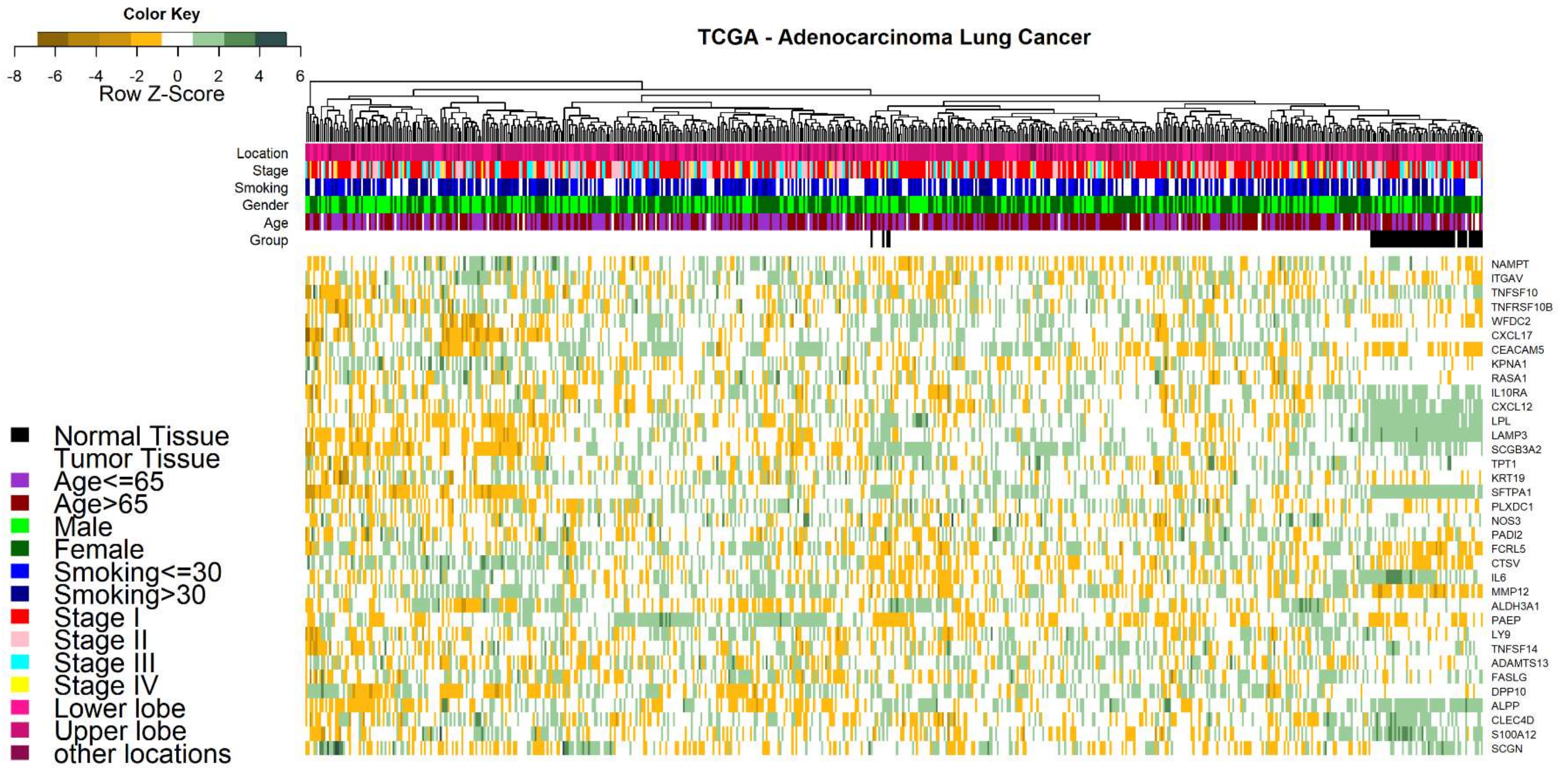

**(B) Squamous lung cancer tissues**

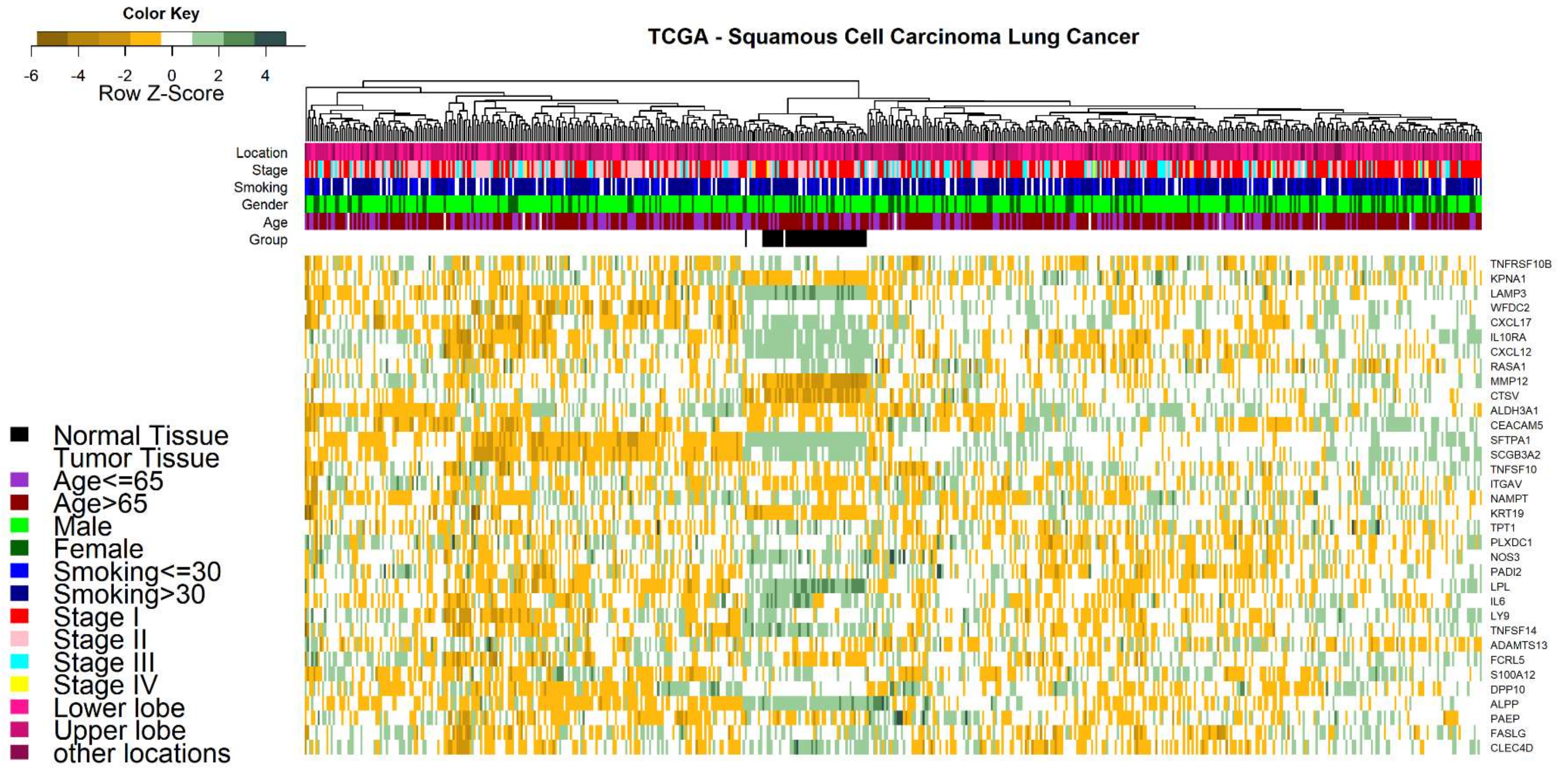

**Supplementary Figure 4: The ROC-AUC of the combined model including Brock score and the top 9 circulating protein markers in comparison to the Brock model, overall and in different subgroups by smoking and nodule size.**

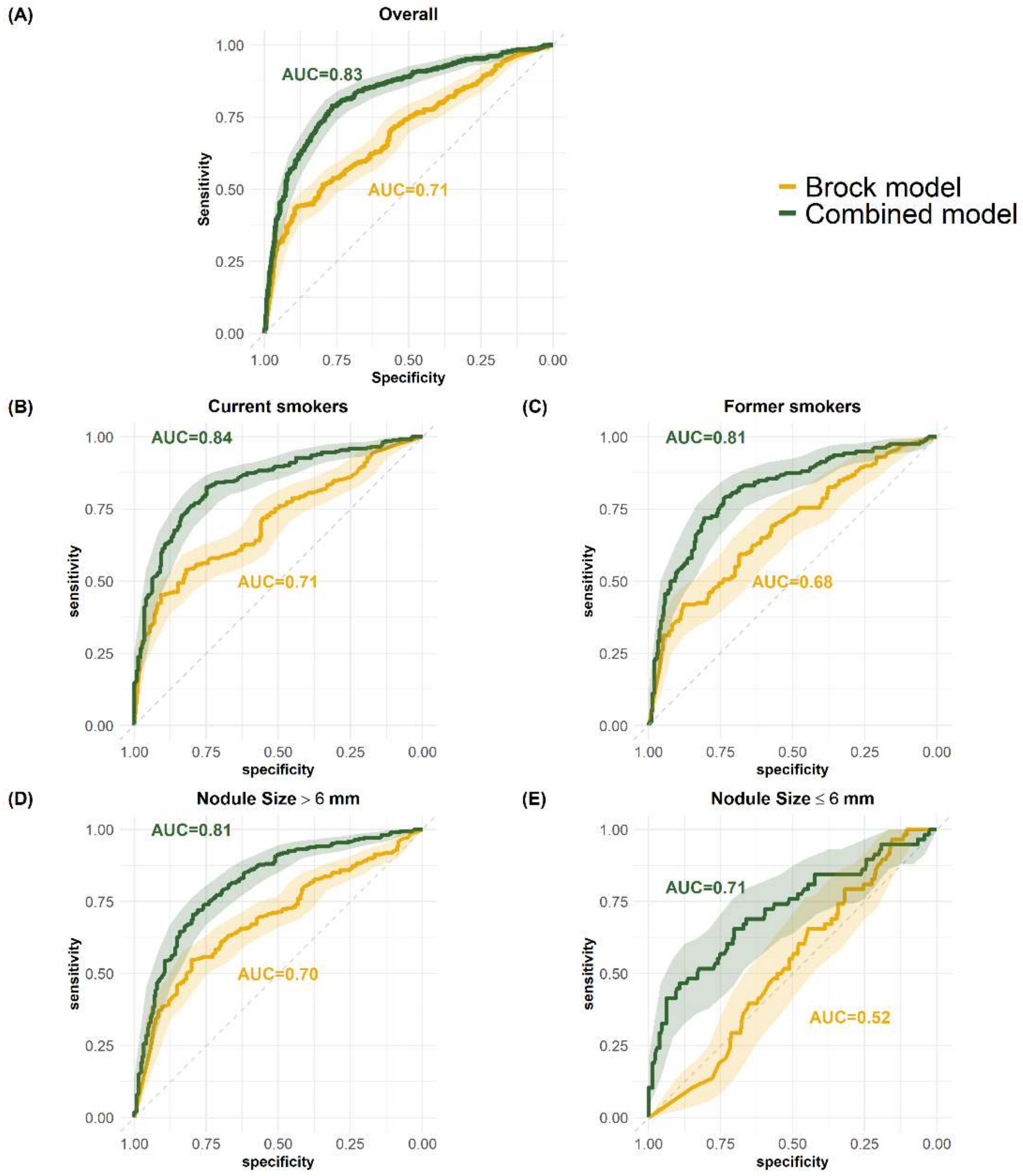

**Supplementary Figure 5: Violin plots of the top circulating protein markers illustrating differential expression of the top protein markers in lung cancer subjects, subjects with benign nodules and subjects with no nodule.**

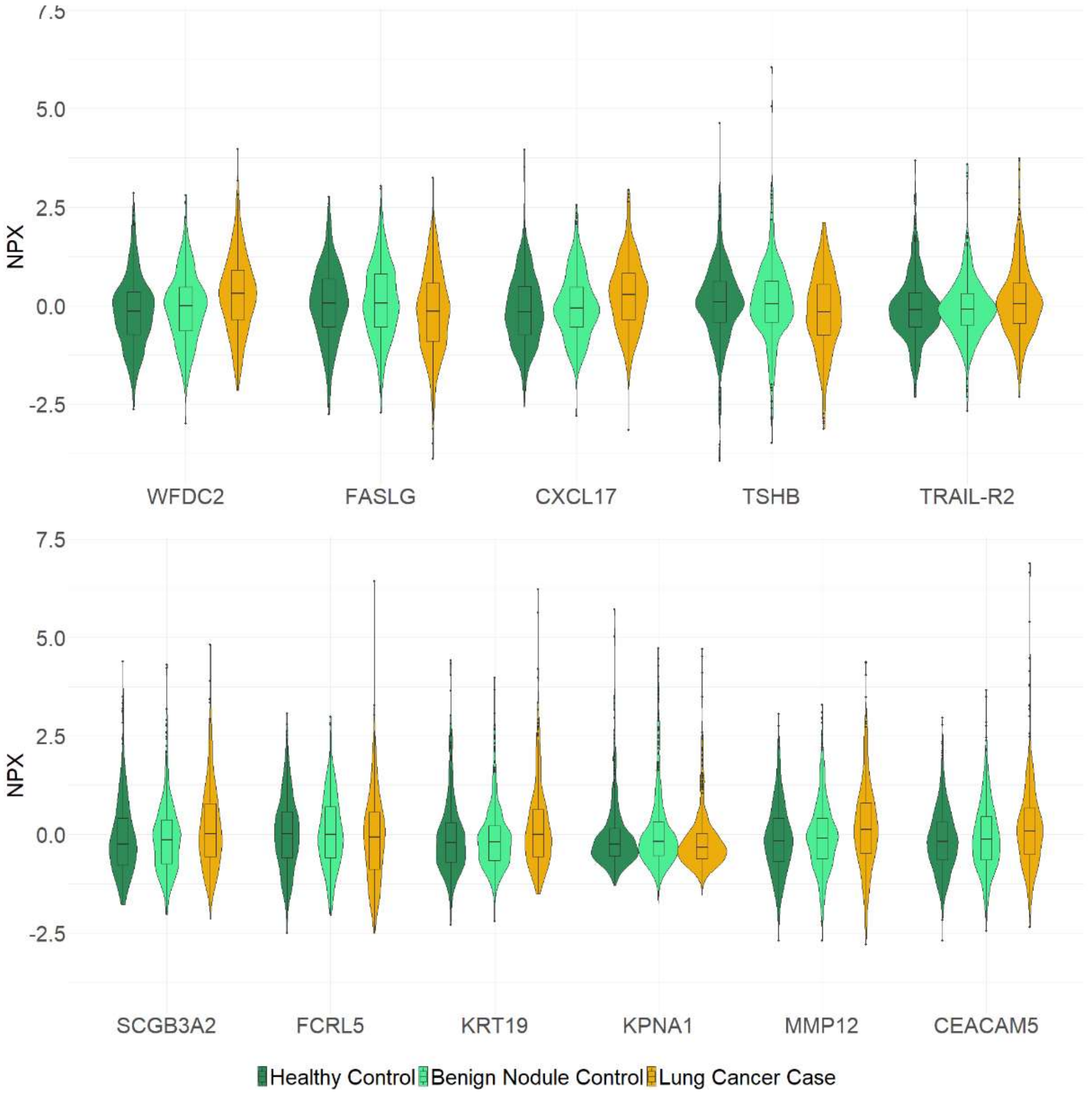
